# A systematic review and meta-analysis of the efficacy and safety of pharmacological treatments for obesity in adults: 2026 Update

**DOI:** 10.64898/2026.04.19.26351196

**Authors:** Andreea Ciudin, Jennifer L. Baker, Andrej Belančić, Luca Busetto, Dror Dicker, Ľubomíra Fábryová, Gema Frühbeck, Gijs H. Goossens, Jason Gordon, Matteo Monami, Paolo Sbraccia, Borja Martinez-Tellez, Volkan Yumuk, Barbara McGowan

**Affiliations:** Andreea Ciudin, Diabetes and Metabolism Research Group, VHIR, Endocrinology Department, Vall d’Hebron University Hospital, Autonomous University Barcelona, Barcelona, Spain; CIBERDEM (Instituto de Salud Carlos III), Madrid, Spain; Jennifer L. Baker, Center for Clinical Research and Prevention, Copenhagen University Hospital-Bispebjerg and Frederiksberg, Frederiksberg, Denmark; Department of Basic and Clinical Pharmacology and Toxicology, University of Rijeka, Faculty of Medicine, Rijeka, Croatia; Department of Medicine, University of Padova, Padova, Italy; Internal Medicine Department and Obesity Clinic, Hasharon Hospital-Rabin Medical Center. Faculty of Medicine, Tel-Aviv University, Tel-Aviv, Israel; Department for diabetes, metabolic diseases and nutritional disorders, Slovak Medical University, Biomedical Center of the Slovak Academy of Sciences, Bratislava, Slovakia; Department of Endocrinology & Nutrition, Institute for Nutrition & Health, Clínica Universidad de Navarra, CIBEROBN - Instituto de Salud Carlos III, IdiSNA, Pamplona, Spain; Department of Human Biology, Institute of Nutrition and Translational Research in Metabolism (NUTRIM), Maastricht University Medical Center+, Maastricht, the Netherlands; Faculty of Medicine & Health Sciences, School of Medicine, University of Nottingham.; Diabetes Unit, Careggi Teaching Hospital and University of Florence, Florence, Italy; Department of Systems Medicine, University of Rome Tor Vergata, Rome, Italy; Department of Nursing, Physiotherapy and Medicine, CIBIS Research Center, University of Almería, Almería, Spain; CIBEROBN, Instituto de Salud Carlos III, Granada, Spain; Division of Endocrinology, Metabolism and Diabetes, Istanbul University-Cerrahpaşa, Cerrahpaşa Medical Faculty, Istanbul, Turkey; Department of Endocrinology and Diabetes, Guys & St Thomas ‘s Hospital NHS Trust, London, UK

## Abstract

This updated systematic review and network meta-analysis evaluated the efficacy and safety of obesity management medications (OMMs) in terms of reducing body weight and obesity-related complications. Medline and Embase were searched up to 21 November 2025 for randomized controlled trials comparing OMMs versus placebo or active comparators in adults. The primary endpoint was percentage total body weight loss (TBWL%) at the end of the study. Secondary endpoints were TBWL% at 1, 2 and ≥3 years, anthropometric, metabolic, mental health and quality-of-life outcomes, cardiovascular morbidity and mortality, remission of obesity-related complications, serious adverse events and all-cause mortality.

Sixty-six RCTs (66 comparisons) were identified – orlistat (22), semaglutide (18), liraglutide (11), tirzepatide (8), naltrexone/bupropion (5) and phentermine/topiramate (2) – enrolling 63,909 patients (34,861 and 29,048 with active compound and placebo, respectively). All OMMs showed significantly greater TBWL% versus placebo; tirzepatide and semaglutide exceeded 10% TBWL and showed the most favourable glycaemic effects. Semaglutide reduced major adverse cardiovascular events and all-cause mortality. In dedicated complication-specific trials, semaglutide and tirzepatide showed benefit on heart-failure-related outcomes; tirzepatide was associated with improved obstructive sleep apnoea syndrome and semaglutide with knee osteoarthritis pain remission. Tirzepatide and semaglutide were associated with improvements in metabolic dysfunction-associated steatohepatitis remission, and semaglutide with improvement in liver fibrosis. No OMMs were associated with an increased risk of serious adverse events. These updated results reinforce the need to individualize OMMs selection according to weight-loss efficacy, complication profile and safety.

## Introduction

The prevalence of overweight and obesity continues to rise globally. In 2022, 2.5 billion adults worldwide were living with overweight, including more than 890 million living with obesity, and projections suggest that the number of adults living with obesity could exceed 1.1billion by 2030 (WHO 2025; World Obesity 2025). Obesity is associated with increased risk of cardiometabolic disease, several cancers, impaired physical functioning and premature mortality, and it imposes a substantial burden on health systems and economies (Nagi 2024). Lifestyle intervention remains the foundation of care, but long-term weight loss and weight-loss maintenance are difficult to achieve for many individuals.

Obesity a recognized as a chronic, relapsing, adiposity-based disease. Reflecting this understanding of its biology, the 2024 European Association for the Study of Obesity (EASO) framework recommends moving beyond body mass index alone toward diagnosis and staging approaches that also incorporate body fat distribution and medical, functional and psychological complications (Busetto 2024). This reframing supports treatment strategies based on disease severity and complication burden, with escalation beyond behavioural intervention when clinically indicated (Ciudin 2026; ADA 2026).

Recent advances in obesity pharmacotherapy have expanded therapeutic options and raised expectations not only for the magnitude of weight loss that can be achieved, but also for improvement in obesity-related complications (Ciudin 2026). Semaglutide 2.4 mg reduced major adverse cardiovascular events in adults with overweight or obesity and established cardiovascular disease without diabetes in the SELECT trial (Lincoff 2023; Kushner 2025). Additional randomised controlled trials (RCTs) have demonstrated benefit of semaglutide in the obesity phenotype of heart failure with preserved ejection fraction (Kosiborod 2023; Kosiborod 2024). Tirzepatide, a dual glucagon-like peptide-1 (GLP-1) receptor agonist and glucose-dependent insulinotropic polypeptide (GIP) receptor agonist, has demonstrated substantial weight-loss efficacy alongside efficacy across multiple complication-specific RCTs, including heart-failure-related outcomes (Packer 2025; Borlaug 2025), moderate-to-severe obstructive sleep apnoea (Malhotra 2024; Kanu 2025), and metabolic dysfunction-associated steatohepatitis (Loomba 2024). Both semaglutide and tirzepatide have been shown to increase metabolic dysfunction-associated steatohepatitis (MASH) resolution (Sanyal 2025; Loomba 2024), whereas improvement in liver fibrosis has been demonstrated for tirzepatide (Loomba 2024). More recently, the SURMOUNT-5 trial provided direct comparative evidence between tirzepatide and semaglutide (Aronne 2025).

However, direct head-to-head evidence remains limited, and RCTs differ in eligibility criteria, follow-up duration and the obesity-related outcomes assessed. The use of network meta-analysis (NMA) can help address these gaps by integrating direct and indirect evidence across available agents. A previous systematic review and NMA synthesized evidence available to 31 January 2025 and informed development of the first GRADE-based EASO algorithm for pharmacological obesity treatment (McGowan 2025a; McGowan 2025b). Since then, additional RCTs meeting the study inclusion criteria have become available (n=14), including new active-comparator and complication-specific studies. We therefore conducted an updated systematic review and NMA of European Medicines Agency (EMA) approved obesity management medications (OMMs) available in Europe, incorporating evidence published up to 21 November 2025 to reassess their comparative efficacy and safety for body weight reduction and obesity-related complications in adults.

## Methods

This NMA is reported in accordance with the Preferred Reporting Items for Systematic Reviews and Meta-Analyses (PRISMA) statement (Supplementary Table 1).

### Study search and selection

The protocol for this meta-analysis and NMA was registered in PROSPERO (registration number: CRD42024625338) and published in a previous article (McGowan 2025a). The present analysis included all placebo- or active-controlled RCTs that enrolled adults (≥18 years) with obesity (body mass index (BMI) ≥30 kg/m²) or overweight (BMI >25 kg/m²), with a treatment duration of at least 48 weeks, and that compared OMMs indicated for obesity treatment with either placebo or another OMM. Furthermore, the RCTs needed to be performed on OMMs approved by the EMA and available in at least one EASO member country as of 21 November 2025. The OMMs and doses included were as follows: orlistat (360 mg), phentermine plus topiramate (15/92 mg), naltrexone plus bupropion (32/360 mg), liraglutide (3.0 mg), semaglutide (2.4 mg) and tirzepatide (10-15 mg).

To update and extend the previous review, a Medline and Embase search was performed up to 21 November 2025 using the validated search strings below, with findings integrated with the previously identified evidence base (Supplementary Fig. 1). Duplicate records were removed with EndNote X9 (Clarivate Analytics, Philadelphia, PA, USA).

### Data extraction

Information on baseline characteristics and outcomes was extracted, including age, gender, proportion of patients with type 2 diabetes (T2D), baseline BMI, percentage total body weight loss (TBWL%), waist circumference, body composition, proportion of patients achieving at least 5%, 10%, 15%, 20% and 25% body weight reduction, remission or improvement/resolution of obesity-related complications, serious adverse events (SAE), mortality, major adverse cardiovascular events (MACE), fasting plasma glucose (FPG), HbA1c, lipid profile, estimated glomerular filtration rate (eGFR), creatinine, albuminuria, mental health parameters and quality of life (QoL). Two authors performed data extraction independently, and disagreements were resolved by a third investigator.

### Quality assessment

The risk of bias was assessed using the Cochrane-recommended tool to determine the risk of bias in RCTs (Higgins 2011). The risk of bias was described and evaluated in seven specific domains: random sequence generation, allocation concealment, blinding of participants and personnel, blinding of outcome assessment, incomplete outcome data, selective reporting and other biases. The results of these domains were graded as low, high or unclear risk of bias. Two researchers independently assessed the risk of bias in individual studies, with discrepancies resolved by a third researcher.

### Outcomes

#### All included trials

The principal endpoint was TBWL%; secondary endpoints evaluated at the end of the trial period were:

a. TBWL% at 52 weeks, 53-104 weeks, 105-156 weeks and >156 weeks
b. Change in end-point BMI and waist circumference
c. Change in total fat mass, subcutaneous fat mass and visceral fat mass
d. Change in fat-free mass
e. The proportion of patients achieving at least 5%, 10%, 15%, 20% and 25% body weight reduction
f. T2D, hypertension and dyslipidemia remission. T2D remission was defined as HbA1c <6.5% at end point
g. MASH resolution (defined as no steatotic liver disease without worsening of fibrosis), improvement of liver fibrosis (defined as a decrease of at least one fibrosis stage without worsening of MASH) (Loomba 2023; Loomba 2024), OSAS resolution (defined as AHI <5 or AHI of 5 to 14) (Malhotra 2024), and improvement of KOA (improvement in pain/physical functioning items of any validated scale assessing QoL in patients with KOA)
h. Reduction in hospitalisation for heart failure (HHF), considering only studies in which these events were formally adjudicated
i. Any SAE
j. All-cause mortality
k. MACE (composite of non-fatal myocardial infarction, non-fatal stroke and cardiovascular mortality), considering only studies in which these events were formally adjudicated
l. End-point FPG, HbA1c, lipid profile, eGFR, creatinine and albuminuria
m. Change in mental health parameters
n. QoL

#### Trials performed in populations with pre-existing comorbid conditions

Trials performed in specific populations of subjects with obesity and other comorbid conditions were also analysed separately. For all the above-reported conditions only adjudicated events were considered.

- Established CVD: Primary endpoint: incidence reduction of MACE. Other critical endpoints: body weight reduction (TBWL%), all-cause and cardiovascular mortality reduction.
- Heart failure: Primary endpoint: reduction of HHF. Other critical endpoints: body weight reduction (TBWL%), incidence of MACE, improvement of Kansas City cardiomyopathy questionnaire (KCCQ) clinical summary score, change in 6-minute walking test distance, and all-cause and cardiovascular mortality reduction.
- Pre-diabetes: Primary endpoint: normoglycemia restoration. Other critical endpoints: body weight reduction (TBWL%); lipid and blood pressure profile, renal function, reduction of incident T2D, and improvement of metabolic control (HbA1c and FPG).
- Type 2 diabetes: Primary endpoint: complete T2D remission. Other critical endpoints: body weight reduction (TBWL%); lipid and blood pressure profile, renal function, and improvement of metabolic control (HbA1c and FPG).
- MASLD: Primary endpoint: MASH remission. Other critical endpoints: body weight reduction (TBWL%) and improvement of fibrosis and liver indexes.
- OSAS: Primary endpoint: OSAS remission. Other critical endpoints: body weight reduction (TBWL%) and improvement of parameters evaluating apnea.
- KOA: Primary endpoint: KOA improvement assessed with scales evaluating osteoarthritis outcome scores (Western Ontario and McMaster Universities Osteoarthritis Index [WOMAC] – pain and physical function score). Other critical endpoints: body weight reduction (TBWL%), improvement of 6-m walking distance, and opioids use.

### Statistical analyses

Analyses were conducted in accordance with the previously described methodology (McGowan 2025a). NMAs were performed for outcomes reported by 10 or more RCTs, allowing joint synthesis of direct and indirect evidence within a connected treatment network. Outcomes informed by fewer than 10 RCTs were analysed using conventional pairwise meta-analysis. Where only one RCT was available within a subgroup, findings were presented descriptively, as meta-analysis was considered potentially misleading.

For binary outcomes, treatment effects were expressed as odds ratios (ORs) with 95% confidence intervals (CIs), and for continuous outcomes as weighted mean differences (WMDs) with 95% CIs. All analyses were performed using random-effects models to account for between-study heterogeneity. For studies reporting least-squares means with standard errors or confidence intervals, standard deviations were derived as s.d. = √(number of patients) × (confidence interval upper limit − confidence interval lower limit) / 3.92 and s.d. = √(number of patients) × s.e., respectively.

Prespecified subgroup analyses were conducted according to OMM, BMI category (mean BMI at enrolment <30, 30–34.9, 35–39.9 and >40 kg/m²), T2D (RCTs enrolling at least 75% of participants with diabetes), prediabetes, established CVD, HF, KOA, MASLD and OSAS. An additional subgroup analysis was performed for body weight outcomes only (TBWL%, waist circumference, BMI, and the proportions of patients achieving at least 5%, 10%, 15%, 20% and 25% body weight reduction) in RCTs specifically designed for body weight reduction.

Heterogeneity was assessed using I² statistics. Pairwise meta-analyses were performed using the ‘meta’ package in R (version 4.5.1), and NMAs using the ‘netmeta’ package in R (version 4.5.1). Placebo was specified as the reference treatment. NMAs enabled indirect comparisons when direct evidence was unavailable and combined direct and indirect evidence to generate overall treatment effect estimates. Consistency between direct and indirect evidence was assessed using H statistics; H < 3 was taken to indicate minimal inconsistency.

Risk-of-bias assessments were performed using Review Manager (RevMan), version 5.4 (Copenhagen: The Nordic Cochrane Centre, The Cochrane Collaboration, 2014). The GRADE methodology (Guyatt 2013) was used to assess the quality of the body of evidence for the principal endpoint, using GRADEpro GDT software (GRADEpro Guideline Development Tool, McMaster University, 2015; available from gradepro.org).

## Results

### Retrieved trials

Supplementary Fig. 1 shows the study flow and results of the searches of the Medline and Embase databases. Supplementary Table 1 lists the trials excluded after full-text review (n=47). Sixty-two RCTs (66 comparisons) evaluating orlistat (n=22 comparisons), semaglutide (n=18 comparisons), liraglutide (n=11 comparisons), tirzepatide (n=8 comparisons), naltrexone plus bupropion (n=5 comparisons), and phentermine plus topiramate (n=2 comparisons), enrolling 63,909 patients (34,861 and 29,048 with active compound and placebo, respectively), were analysed. 14 new RCTs were identified; of these, 6 were primary studies, while remaining 8 provided additional data for previously included (secondary studies). All RCTs were placebo-controlled, except for two trials that reported multiple comparisons: liraglutide versus placebo and orlistat (Astrup 2012), and semaglutide versus liraglutide and placebo (Rubino 2022). One further trial was active-controlled and compared semaglutide with tirzepatide; therefore, the overall number of available comparisons was 66. The main characteristics of the included RCTs are reported in Supplementary Table 2. No studies reported a mean baseline body mass index (BMI) <30 kg/m2.

Study quality was heterogeneous (Supplementary Fig. 2). Most included RCTs were double-blind (71%), whereas a minority inadequately reported attrition and/or allocation procedures or blinding of outcome assessors (27%).

Of the 66 comparisons, 63 were versus placebo. All except five (Nissen 2016; Gudbergsen 2021; Karhunen 2000; Richelsen 2007; Swinburn 2005) reported information on TBWL% at the endpoint (n=61 comparisons). Only three active-comparator comparisons between different OMMs were identified: liraglutide versus orlistat, semaglutide versus liraglutide, and tirzepatide versus semaglutide. These showed greater efficacy for liraglutide than orlistat (weighted mean difference (WMD) 3.80% [0.73, 6.87], p=0.020), semaglutide than liraglutide (WMD 9.4% [6.8, 12.0], p<0.001), and tirzepatide than semaglutide (WMD 6.5% [4.9, 8.1], p<0.001).

### Network meta-analysis

#### TBWL% at different timepoints

Figure 1 shows the number of available RCTs for each comparison, and Fig. 2 and Table 1 show the efficacy results for TBWL% at 52 weeks, 53-104 weeks, 105-156 weeks, >156 weeks, at study endpoint.

**Table 1.**
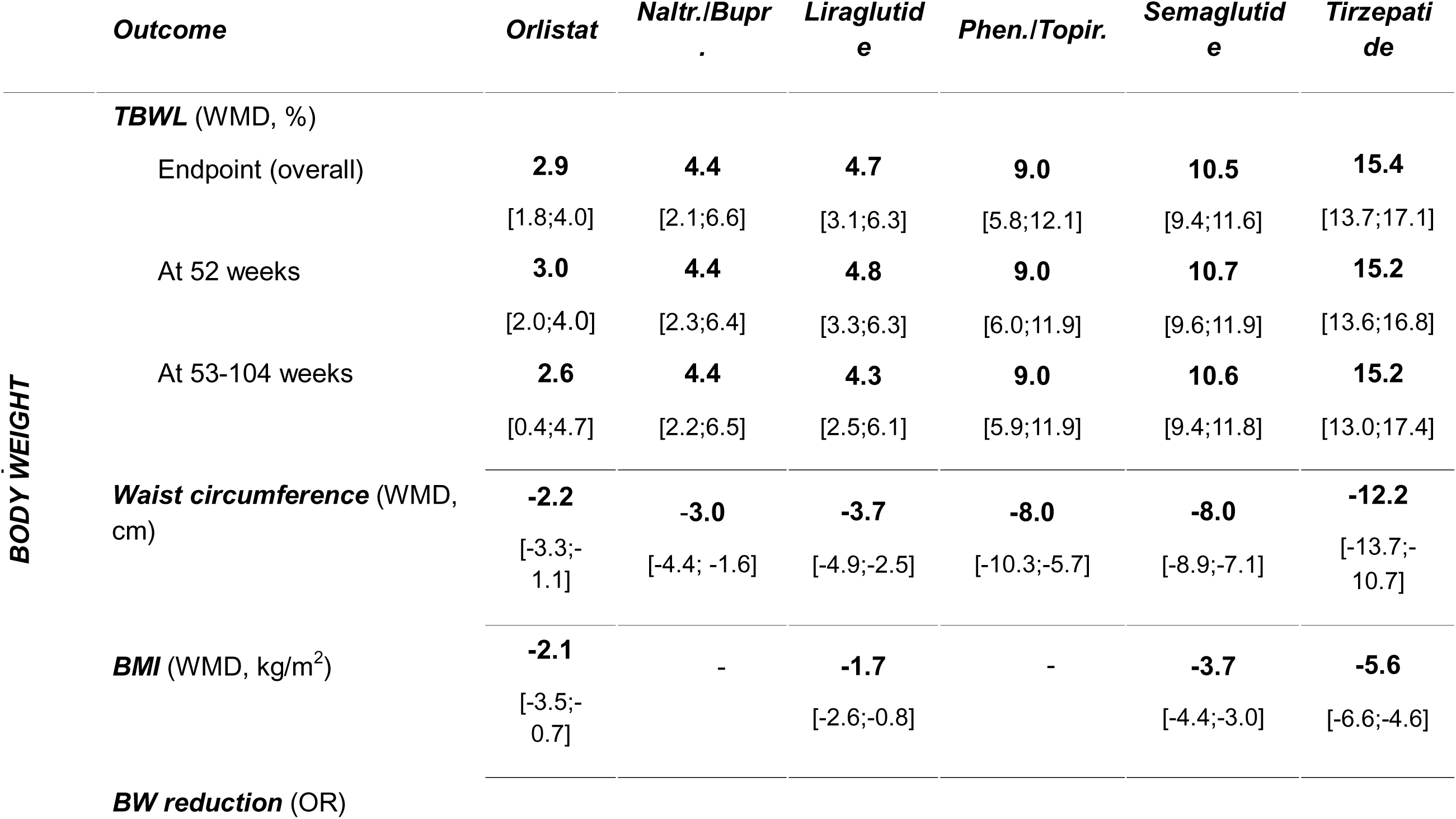
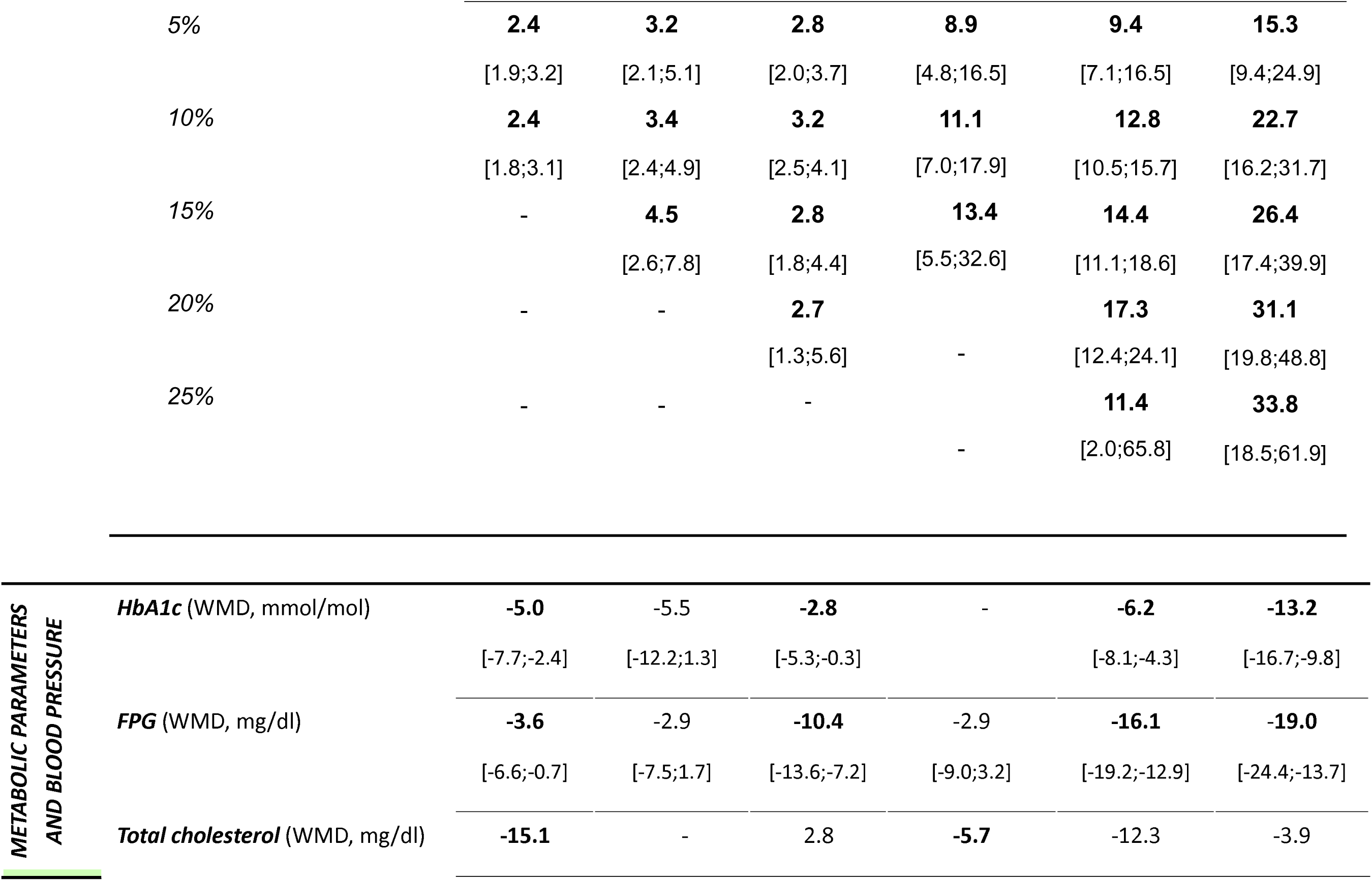
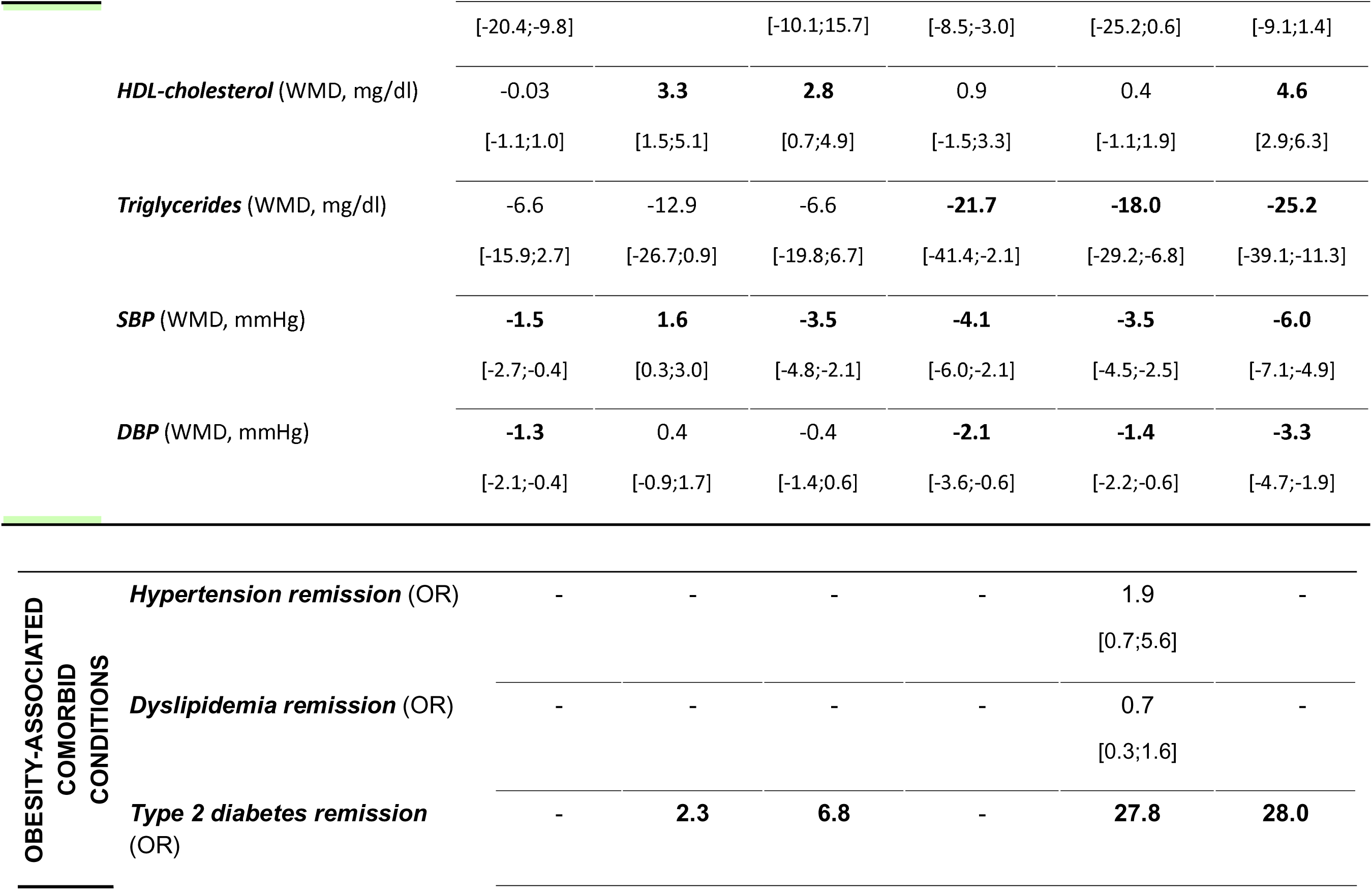
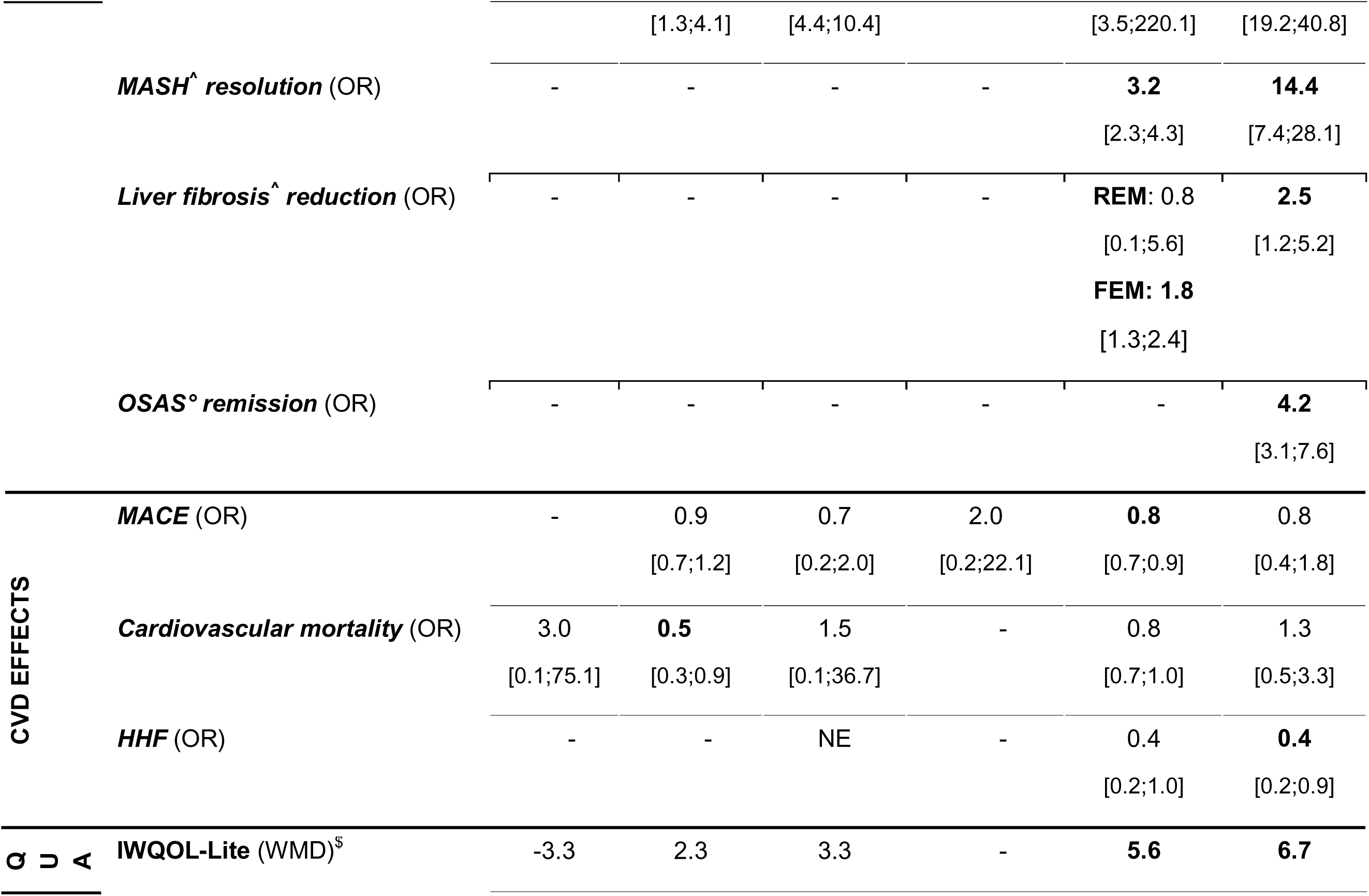
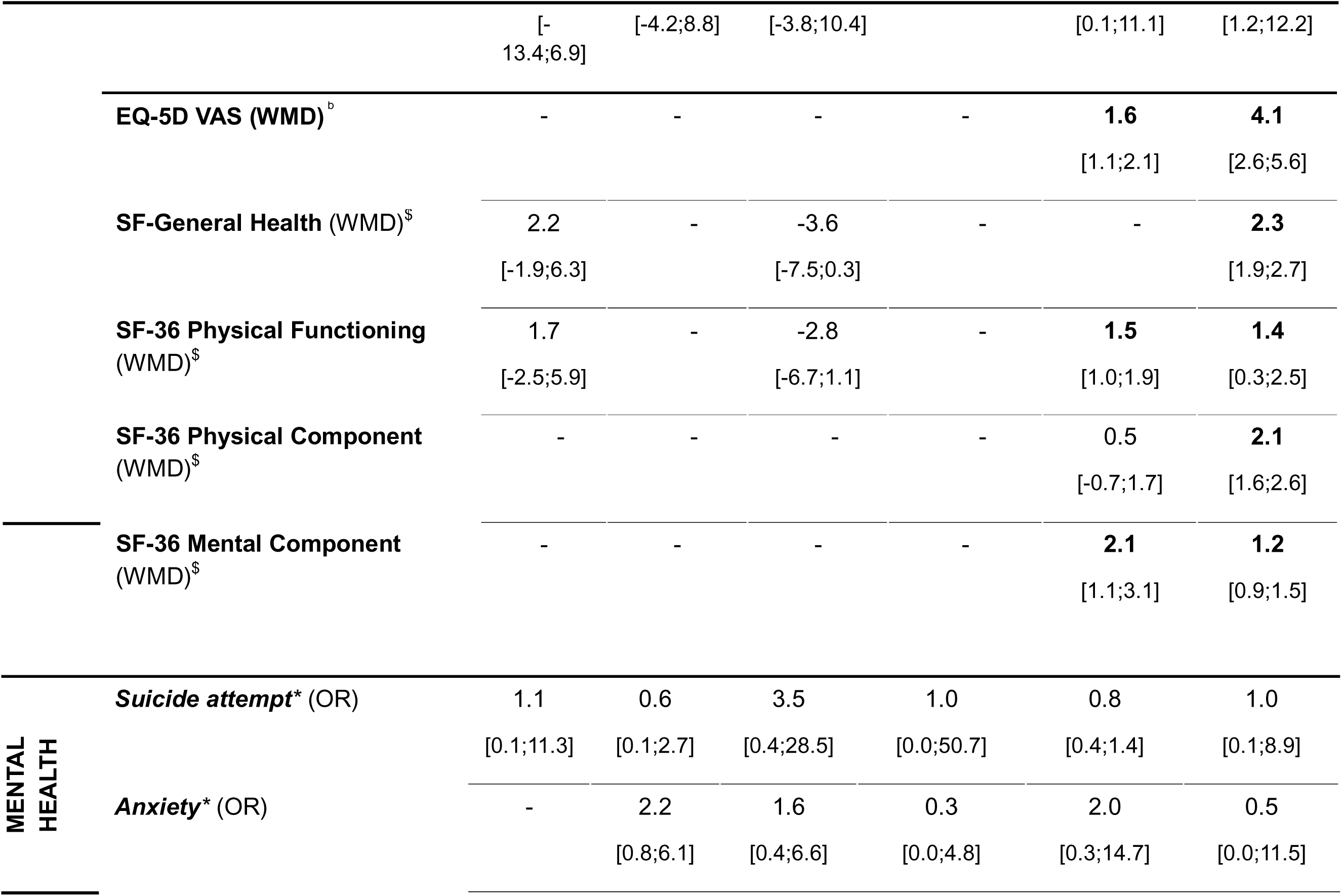
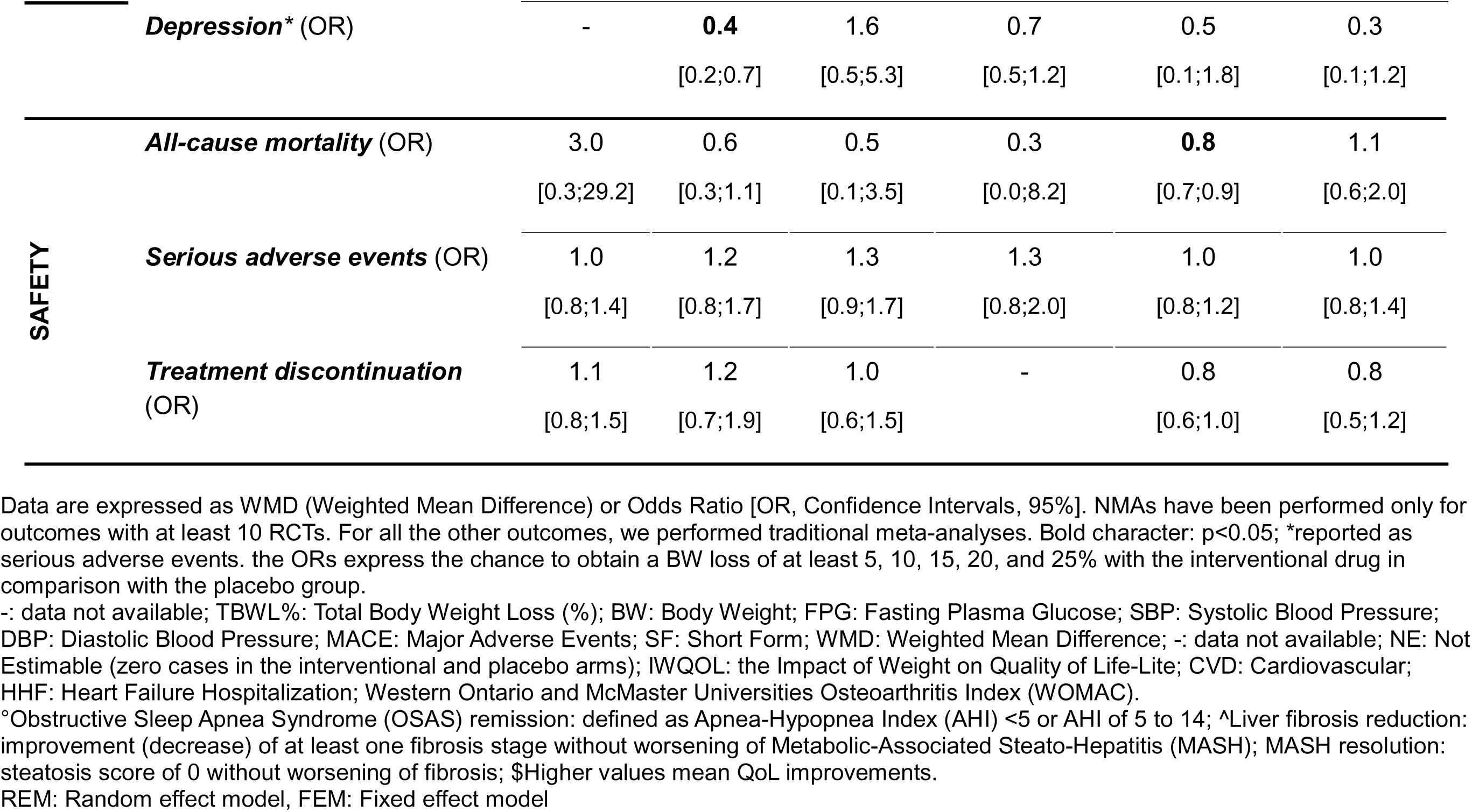
Summary report of NMAs on all critical outcomes at endpoint (placebo-subtracted effect), if not otherwise specified.

Sixty-one comparisons reported information on TBWL% (Fig. 1a). All OMMs were associated with statistically significantly greater TBWL% compared with placebo, with no evidence of inconsistency (H value =1.21; Fig. 2a). The estimated TBWL% was greater than 10% only for tirzepatide and semaglutide (Fig. 2a). Details of direct and indirect estimates are reported in Supplementary Table 3; further visual inspection of funnel plots did not suggest publication bias.

**Figure 1.**
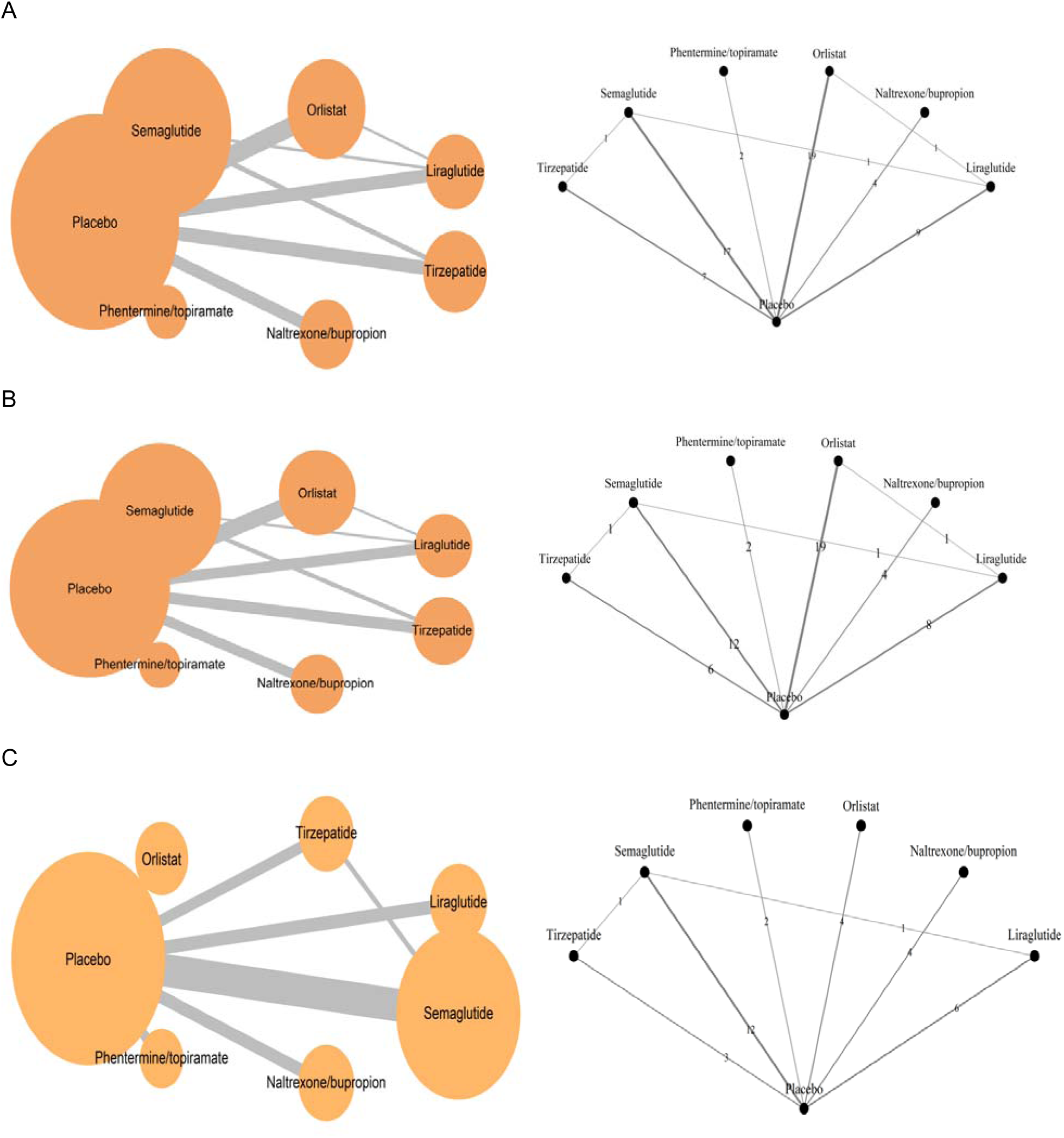
Network plots for trials reporting information on TBWL% at the endpoint (Panel A), 52 weeks (Panel B), and 53-104 weeks (Panel C).

**Figure 2.**
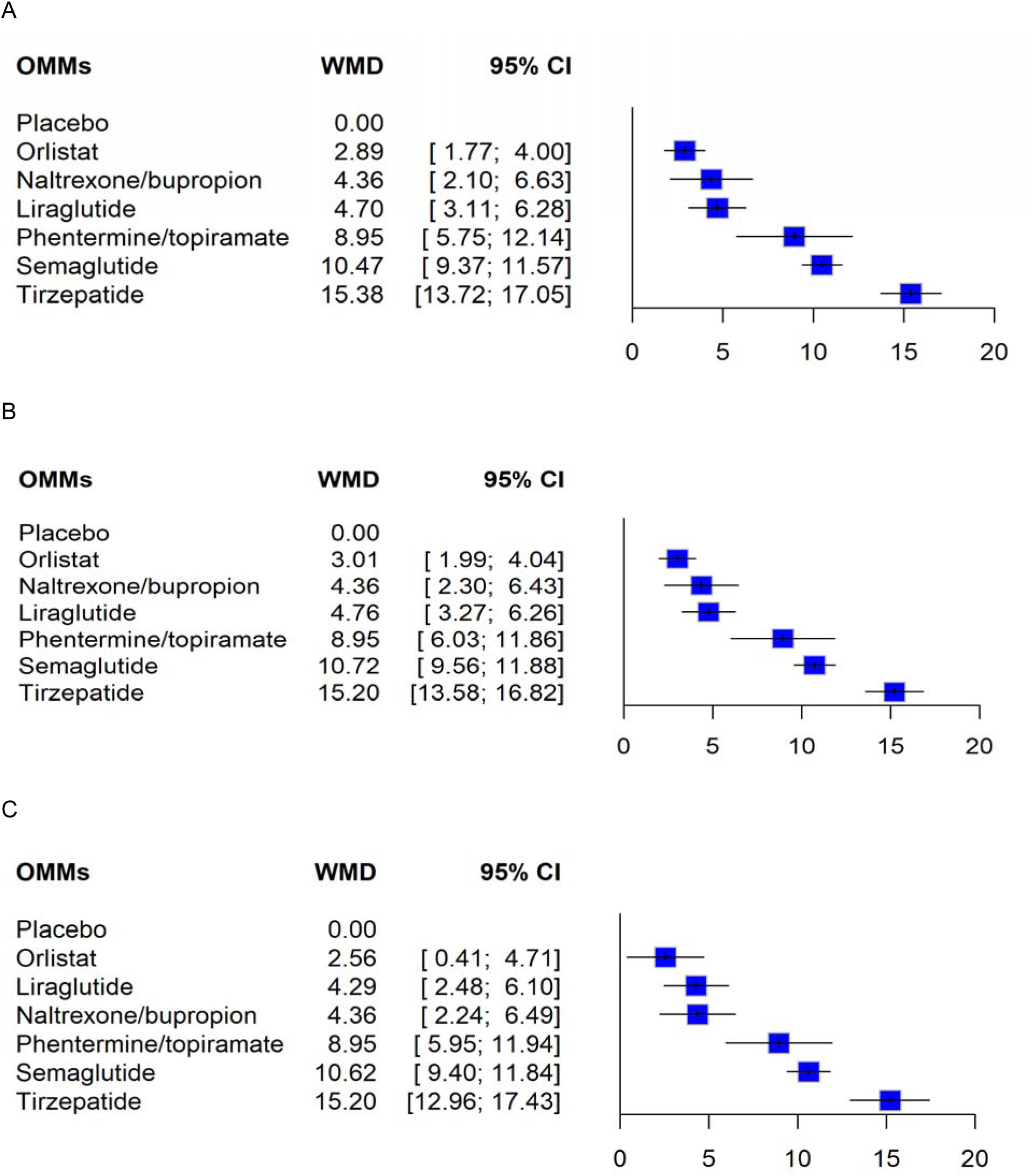
Network forest plots showing the effects of each OMM on TBWL% at the endpoint (Panel A), 52 weeks (Panel B), 53-104 weeks (Panel C)

At 52 weeks (n=54 comparisons; H=1.42), all treatments showed a significantly greater TBWL% versus placebo (Fig. 2b). Similar to the endpoint analyses, semaglutide and tirzepatide achieved a TBWL% of at least 10% (Fig. 2b). Details of direct and indirect estimates are reported in Supplementary Table 4.

Fewer comparisons were available at 53-104 weeks (n=33 comparisons; H=1.43; Fig. 2c and Supplementary Table 5). At 105-156 weeks, data were available from only three trials, one each for liraglutide (Le Roux 2017), orlistat (Torgerson 2004), and semaglutide (Lincoff 2023), reporting placebo-subtracted TBWL of 4.2%, 3.0%, and 8.7%, respectively. Beyond 156 weeks, data were available from one trial with orlistat (Torgerson 2004), reporting 3.0% TBWL versus placebo, and one subgroup analysis of a trial with tirzepatide (Aronne 2024), reporting 19.3% TBWL. For all assessed treatments, the estimated efficacy at endpoint (n=61 comparisons; H=1.21; Fig. 2a) was similar to that observed at 52 weeks and 53-104 weeks.

#### Weight regain after discontinuation of OMMs

Four trials reported information on weight regain after OMMs discontinuation, one each for liraglutide (Le Roux 2017) and tirzepatide (Aronne 2024), and two for semaglutide (Wilding 2022; McGowan 2024). Discontinuation of liraglutide after 12 weeks of treatment (Le Roux 2017) and semaglutide after 26 weeks of treatment (McGowan 2024) was associated, on average, with regain of 47% and 43% of the weight lost at the end of the active treatment period, respectively. Weight regain after discontinuation of semaglutide and tirzepatide after 52 weeks of treatment was 67% and 53% (Wilding 2022; Aronne 2024), respectively.

Waist circumference, BMI, and proportion of patients achieving ≥5-25% TBWL Table 1 and Supplementary Figs. 3 and 4 show the effects of each OMM on endpoint waist circumference and BMI (percentage reduction from baseline). Tirzepatide and semaglutide were associated with greater reductions in both waist circumference and BMI.

The proportions of patients achieving different TBWL% thresholds are reported in Table 1 and Supplementary Figs. 5 and 6. Patients treated with OMMs were more likely to achieve a TBWL of at least 5% than those receiving placebo. A higher degree of TBWL (>20%) was observed only with semaglutide and tirzepatide and, to a lesser extent, with liraglutide. Only tirzepatide was associated with a greater proportion of patients achieving at least 25% TBWL reduction (OR 33.8 [18.4, 61.9], p<0.001). Eight trials (Astrup 2012; Jastreboff 2024; Lundgren 2021; Wilding 2021; Kadowaki 2022; Wharton 2025; Garvey 2025; Kadowaki 2025) reported results on body composition parameters assessed using heterogeneous methods, often in smaller subgroups of included patients. Two studies on semaglutide (Wilding 2021; Kadowaki 2022) did not report any information on dispersion measures and therefore were not included in formal analyses. Semaglutide showed a greater reduction in total fat mass and visceral fat mass, and a greater increase in total lean body mass, compared with placebo (Valera-Mora 2005; Wilding 2021; Kadowaki 2022; Garvey 2025; Wharton 2025). Tirzepatide was associated with a greater reduction in total body fat mass and a significantly lower reduction in total fat-free mass (Loomba 2024), and significantly reduced visceral and subcutaneous fat, as well as hepatic fat, compared with placebo (Kadowaki 2025). Similar results, although to a lesser extent, were observed with liraglutide (Astrup 2012; Lundgren 2021).

When trials were subdivided by baseline BMI (<30, 30-34.9, 35-39.9, and ≥40.0 kg/m2; Table 2), no RCTs on OMMs reported results for patients with overweight. Only two OMMs, semaglutide and phentermine plus topiramate, provided data for class III obesity (BMI ≥40.0 kg/m2; Table 2). Most information on the effects of each included OMM was available for the class II obesity (35-39.9 kg/m2) category, which showed the highest body weight reduction for tirzepatide and semaglutide (TBWL% >10%). A further post-hoc subgroup analysis was performed for RCTs specifically designed for body weight reduction. The results of these analyses, after excluding RCTs with a primary endpoint other than body weight reduction (Nissen 2016; Lincoff 2023; Loomba 2024; Swinburn 2005; Loomba 2023; Malhotra 2024; Packer 2024), are reported in Table 3. Results from these analyses were similar to those of the primary endpoint analysis.

**Table 2.**
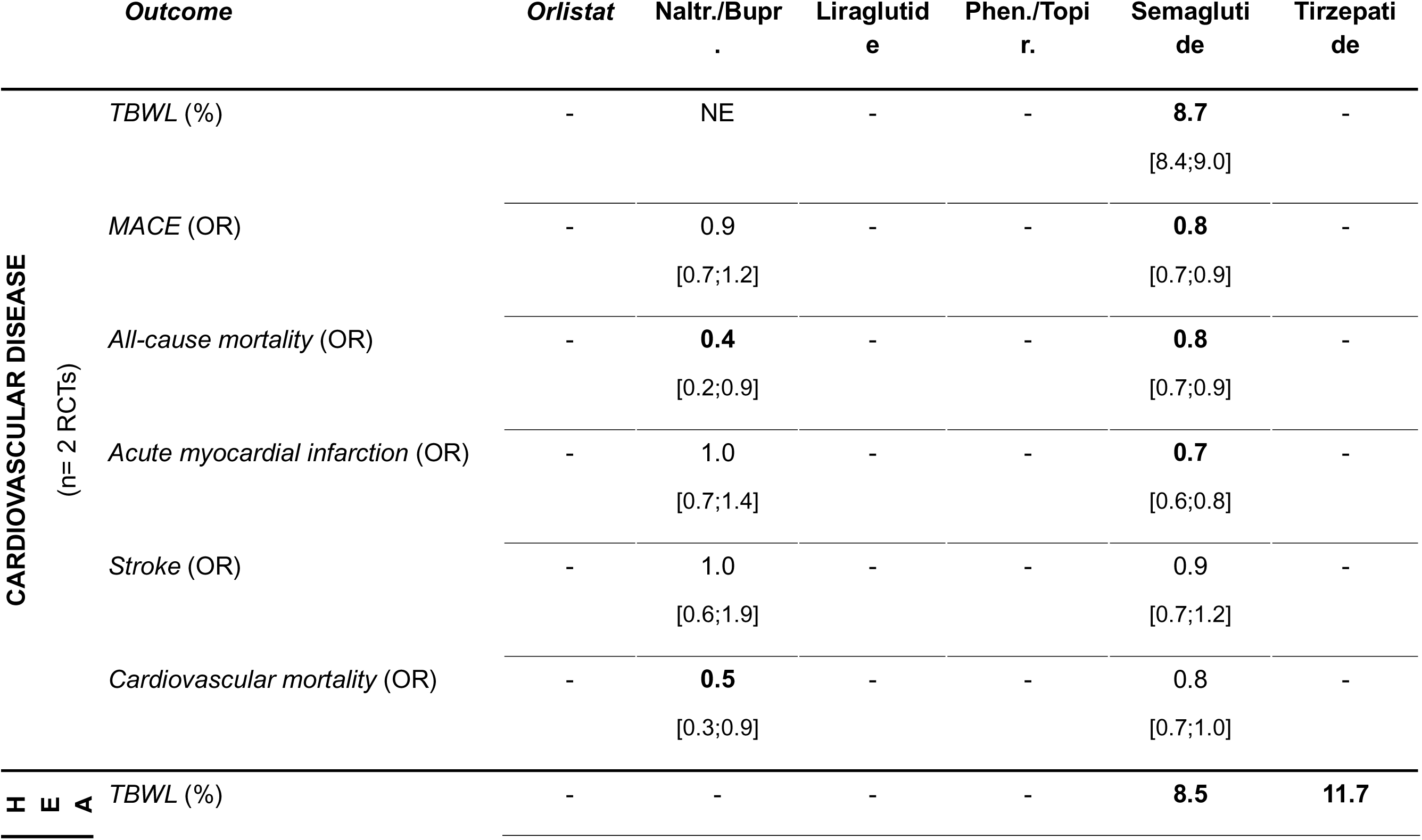
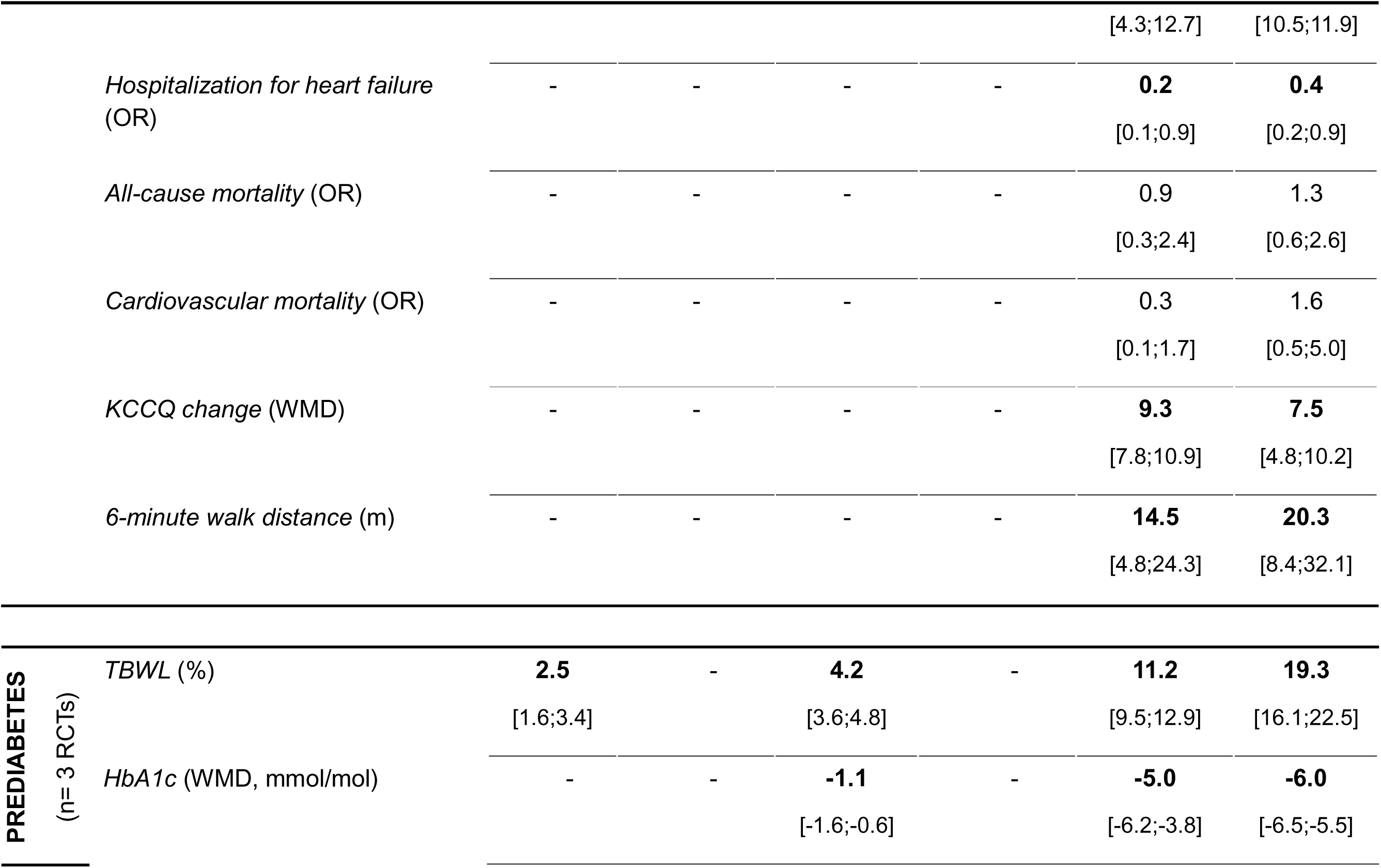
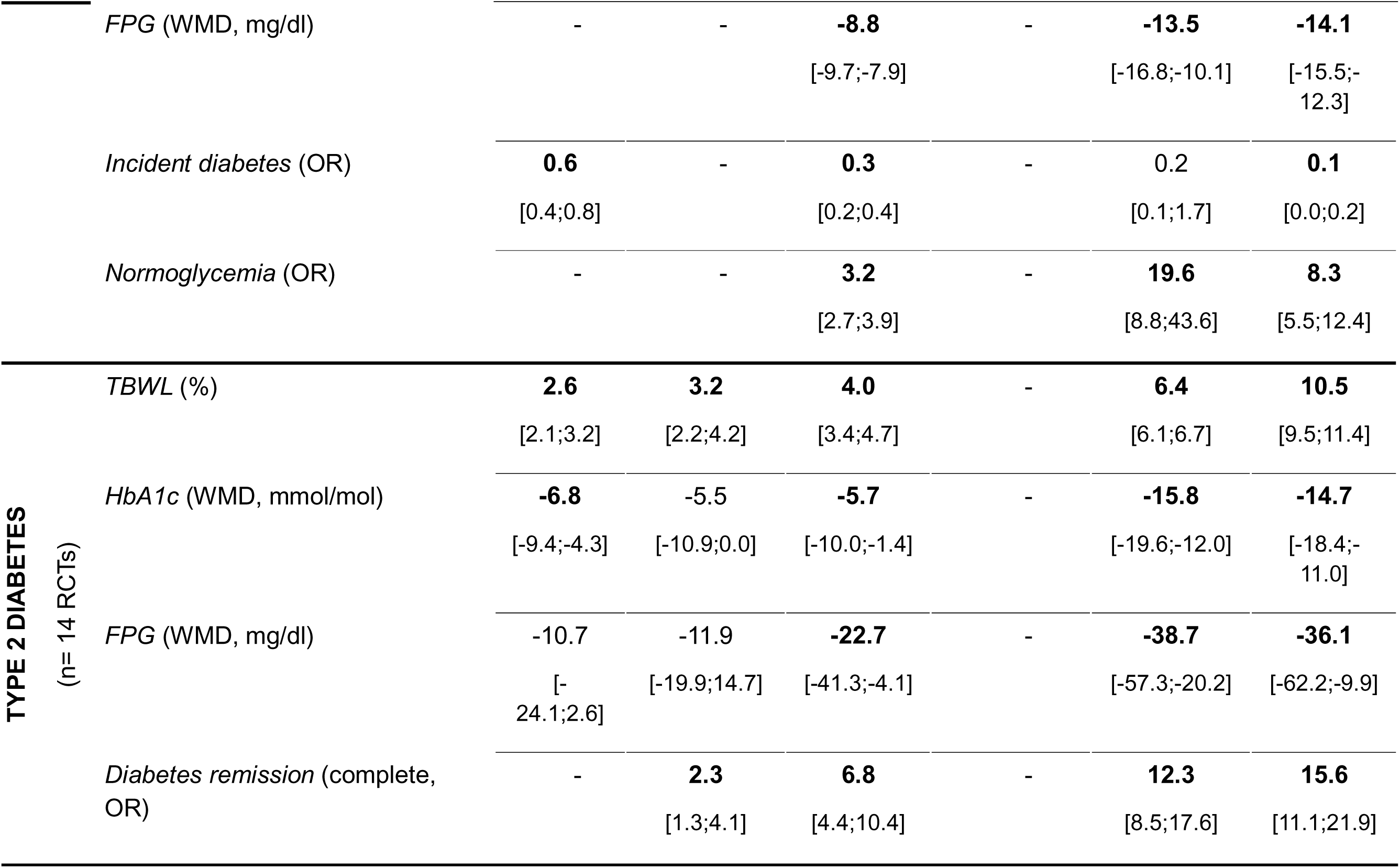
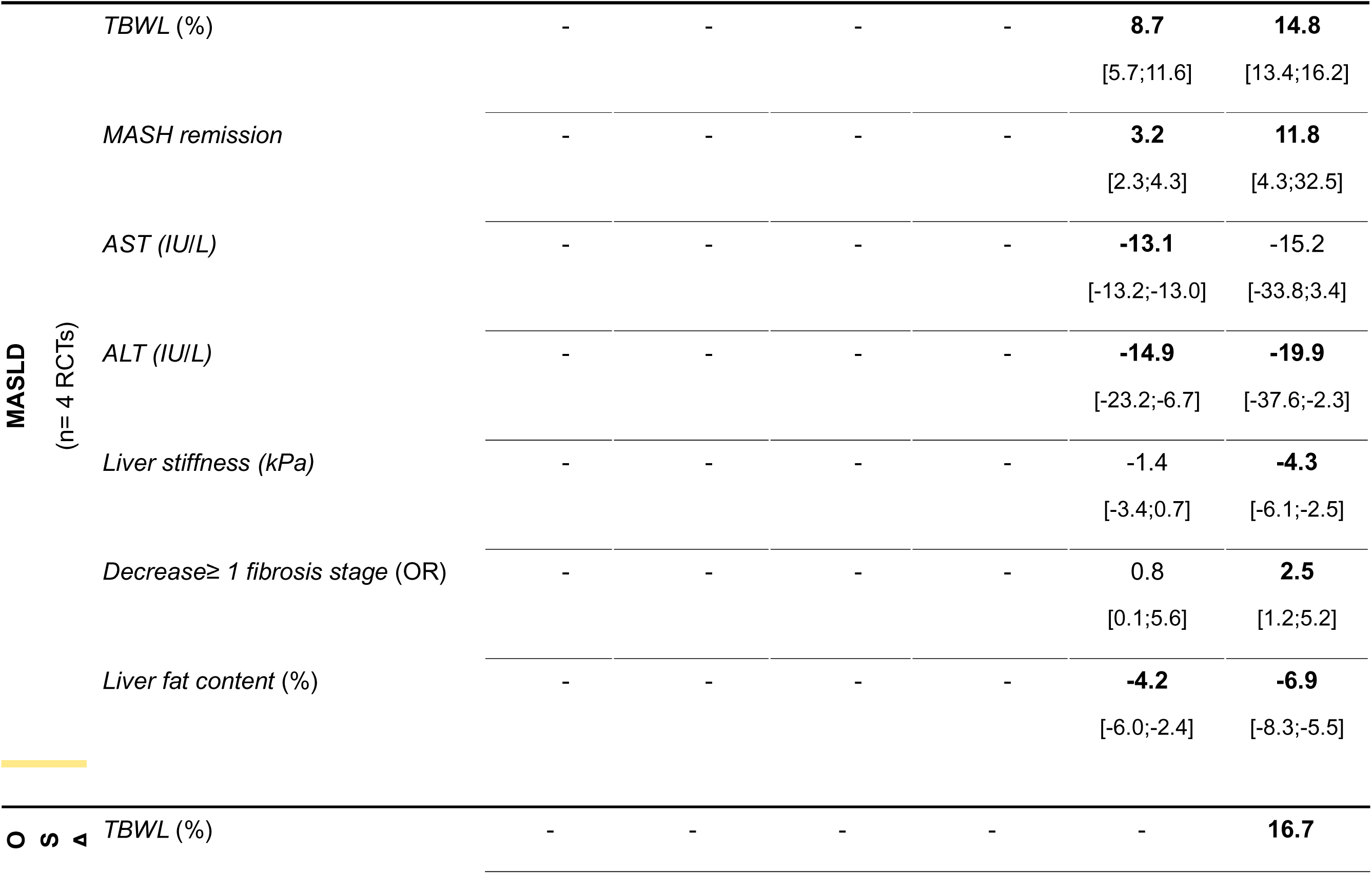
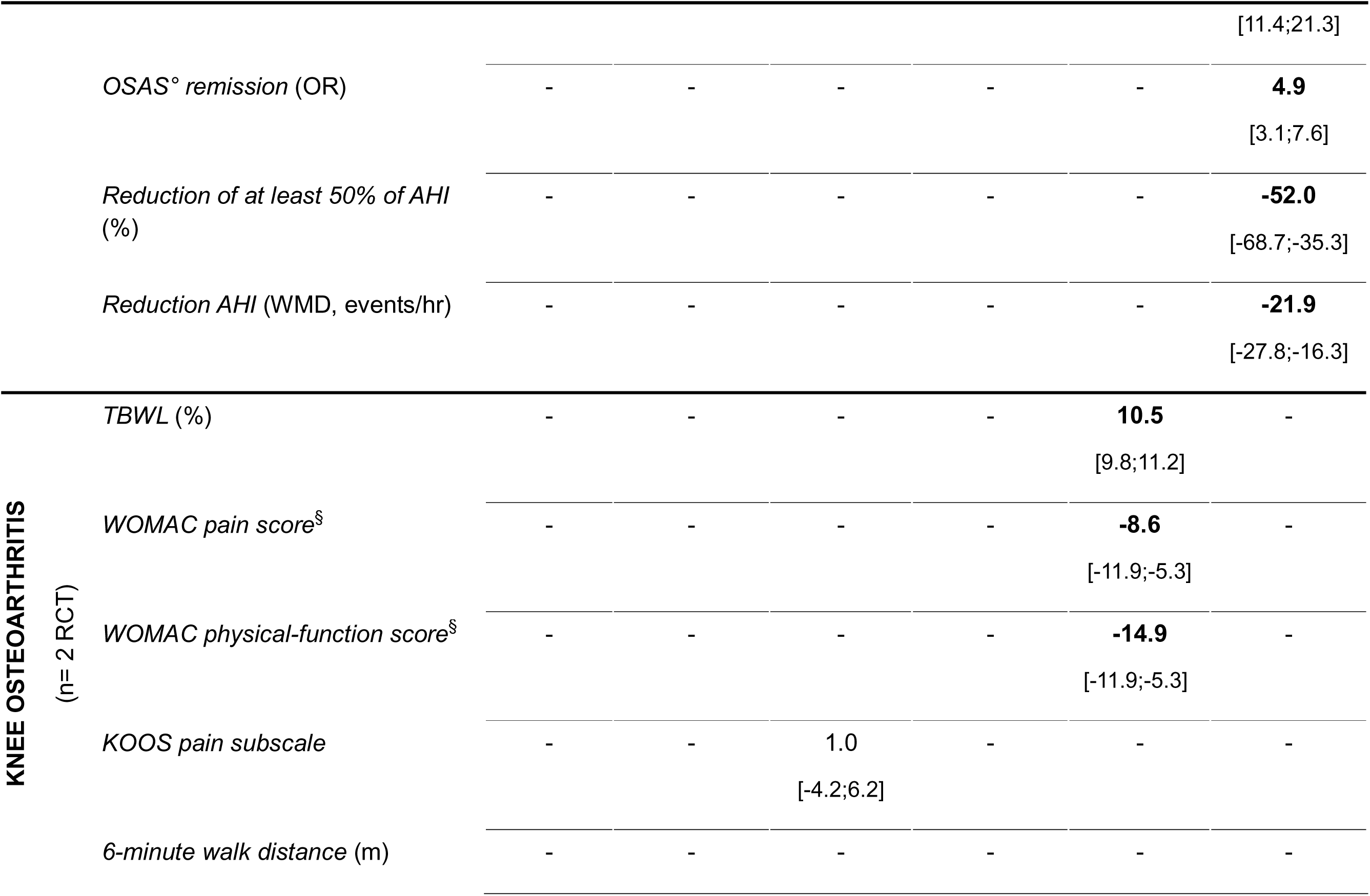
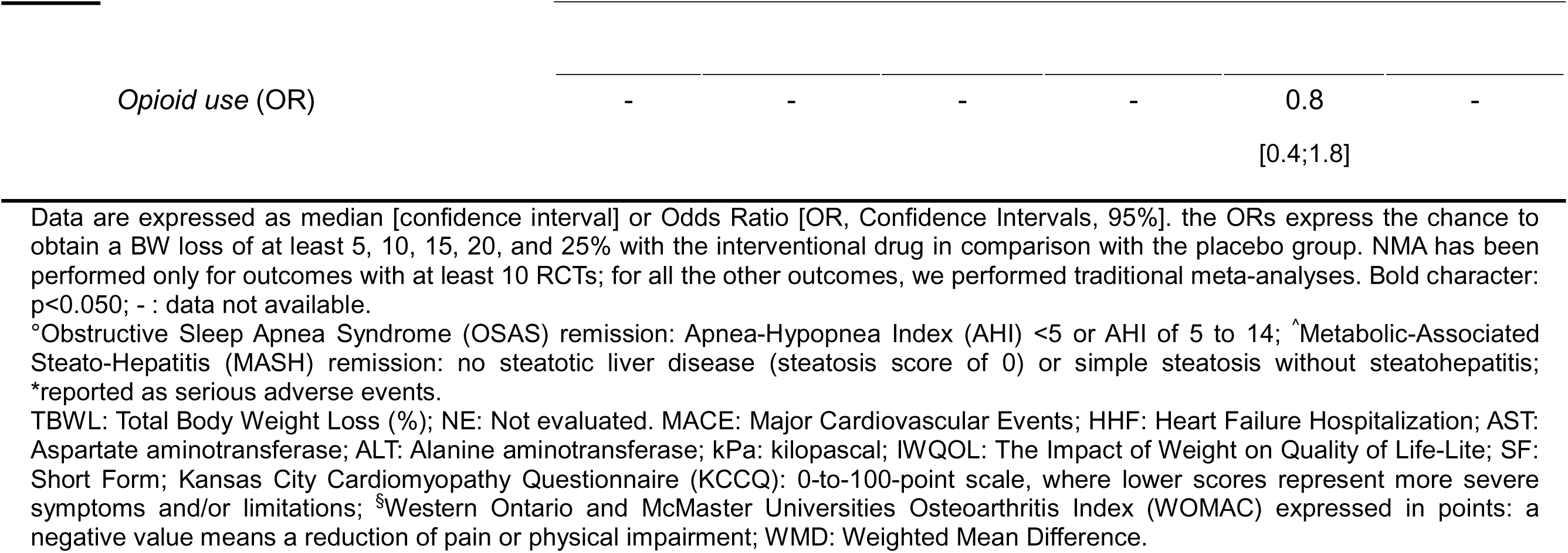
Summary report of OMM effects on critical outcomes at the endpoint for different subpopulations of patients.

**Table 3.** Effects of OMM on endpoint Total Body Weight Loss (TBWL%; placebo-subtracted effect) for different baseline BMI categories.

|  | Endpoint TBWL% |  |  |  |
| --- | --- | --- | --- | --- |
| **BMI** (kg/m<sup>2</sup>) | **<30** | **30-34.9** | **35.0-39.9** | **≥40\*** |
| *Orlistat* | - | **2.3**<br>[1.2;3.4] | **3.2**<br>[1.6;4.8] | - |
| *Naltrexone/Bupropion* | - | - | **4.4**<br>[1.7;7.0] | - |
| *Liraglutide* | - | **5.8**<br>[3.8;7.9] | **4.3**<br>[2.2.;6.4] | - |
| *Phentermine/Topiramate* | - | - | **8.6**<br>[3.4;13.8] | **9.3**<br>[8.3;10.3] |
| *Semaglutide* | - | **13.4**<br>[11.7;15.1] | **10.2**<br>[8.5;11.9] | **10.8**<br>[10.0;11.5] |
| *Tirzepatide* | - | **18.3**<br>[16.4;20.2] | **14.7**<br>[11.6;17.8] | - |

\*Data derived from traditional meta-analysis versus placebo. Data are expressed as median[confidence intervals]; -: data not available. Bold character: p<0.05 vs placebo; BMI: Body Mass Index; TBWL: Total Body Weight Loss (%).

#### Effects on glycaemic control, blood pressure, and lipid profile

Data were analysed across all RCTs reporting each metabolic endpoint, irrespective of the proportion of patients with type 2 diabetes or prediabetes. Table 1 reports results for metabolic parameters (n=29, 36, 31, 37, and 35 RCTs for HbA1c, FPG, total cholesterol, HDL-cholesterol, and triglycerides, respectively) and blood pressure (n=41 and 34 RCTs for systolic and diastolic blood pressure, respectively). Tirzepatide produced a significantly greater reduction in HbA1c than other OMMs, whereas tirzepatide and semaglutide were associated with greater reductions in FPG than the other OMMs (Table 1). T2D remission was reported in only a small number of RCTs (n=5); therefore, traditional meta-analyses were performed for semaglutide only, and estimates from the other three studies were shown in the same figures for convenience (that is, naltrexone plus bupropion, liraglutide, semaglutide, and tirzepatide; Table 1). All studies reported significant results (Table 1). Nine trials reported data on incident T2D; meta-analyses were possible only for orlistat and semaglutide, whereas for phentermine plus topiramate and liraglutide only estimates derived from the original publication were reported (Table 1). All OMMs except orlistat showed significant results (Table 1). Semaglutide and tirzepatide (based on one trial), but not liraglutide, were associated with a higher likelihood of restoring normoglycaemia in patients with pre-existing glycaemic alterations (Table 1).

Orlistat was associated with the greatest reduction in total cholesterol, and statistically significant results versus placebo were also obtained for phentermine plus topiramate. Tirzepatide, naltrexone plus bupropion, and liraglutide were associated with a greater increase in HDL-cholesterol, whereas tirzepatide, semaglutide, and phentermine plus topiramate resulted in a significant reduction in circulating triglyceride levels (Table 1). Table 1 also shows the effects of each OMMs on blood pressure, with naltrexone plus bupropion associated with a significant increase in systolic, but not diastolic, blood pressure. All other OMMs exerted significant favourable effects on systolic blood pressure, and orlistat, phentermine plus topiramate, semaglutide, and tirzepatide on diastolic blood pressure (Table 1). Only a few RCTs reported data on hypertension and dyslipidaemia remission, with clinically inconsequential effects (Table 1).

#### Effects on cardiovascular morbidity and mortality

In total, 34, 38, and 7 RCTs reported information on externally adjudicated MACE, cardiovascular mortality, and HHF, respectively (Supplementary Figs. 7-9). Table 1 and Supplementary Figs. 7-9 show results for these endpoints. Only semaglutide was associated with a significantly lower risk of MACE, although all OMMs were associated with odds ratios below 1 except phentermine plus topiramate (Supplementary Fig. 8b). No information on cardiovascular mortality was reported for phentermine plus topiramate. Semaglutide and naltrexone plus bupropion were associated with a lower risk of cardiovascular mortality (Supplementary Fig. 8c). HHF was significantly reduced by tirzepatide (n=2 RCTs), whereas semaglutide (n=4 RCTs) showed a non-significant trend toward reduction. Liraglutide reported zero events in both arms and therefore the overall risk was not estimable (Supplementary Fig. 9).

#### Effects on quality of life and mental health

A limited number of trials reported data on QoL, using different scales (n=10, 3, 9, and 2 for Impact of Weight on Quality of Life-Lite (IWQOL-Lite), Short Form-36 General Health, Short Form-36 Physical Role Functioning, and Short Form-36 Physical Components, respectively). NMA was feasible only for IWQOL-Lite. The paucity of data for other QoL outcomes did not support NMA, and only traditional meta-analyses were possible when at least two studies were available; for some outcomes, individual studies are presented (Table 1). Naltrexone plus bupropion, liraglutide, tirzepatide, and semaglutide were associated with an improvement in IWQOL versus placebo (Table 1). Only one active-controlled trial (liraglutide versus orlistat) reported data on IWQOL, showing better scores for liraglutide (Astrup 2012). No other significant differences were observed across the remaining scales, except for semaglutide and tirzepatide, which showed improved scores in the Short Form-36 Physical Functioning domain (Table 1).

Suicide attempts and major depression were reported as SAEs, without external adjudication, in a large number of RCTs (Table 1). No significant association was observed between treatment with any OMM and the risk of either suicide attempt or depression, except for naltrexone plus bupropion, which was associated with a significantly lower risk of depression (Table 1). Few studies reported on anxiety (n=11); although an NMA was possible, no significant associations were found for any OMM (Table 1).

#### Safety outcomes

Results for all-cause mortality are reported in Table 1 and Supplementary Fig. 8. Semaglutide was associated with a lower risk of all-cause mortality; the other OMMs did not show any significant effect on this endpoint (Supplementary Fig. 8a). SAEs (Table 1) were reported in most included studies, and no OMM was significantly associated with an increased risk of SAEs compared with placebo. The treatment discontinuation rate was similar to that observed in placebo arms, with no significant between-group differences (Table 1).

#### GRADE evaluation for the primary endpoint

The quality of evidence for the primary endpoint (that is, TBWL%) was rated as high for each OMM (Supplementary Table 6).

#### Subgroup analyses in patients with pre-existing conditions

The effects of each OMM on several prespecified outcomes in specifically designed RCTs or in reported subgroups of patients affected by that comorbid condition were evaluated.

#### Established CVD

Only two trials, one with semaglutide (Lincoff 2023) and one with naltrexone plus bupropion (Nissen 2016), were designed to explore cardiovascular safety in patients with obesity.

Information on TBWL% was available only for semaglutide, with results similar to those obtained in non-cardiovascular outcome trials (8.7%; Table 4). Semaglutide, but not naltrexone plus bupropion, was associated with a significantly lower risk of MACE and acute myocardial infarction in patients with established CVD, whereas cardiovascular mortality was reduced by naltrexone plus bupropion but not by semaglutide. By contrast, both OMMs reported a significantly lower risk of all-cause mortality compared with placebo (Table 4). The GRADE evaluation of the evidence retrieved for the primary endpoint (MACE, Supplementary Table 3) was rated as high and moderate for semaglutide and naltrexone plus bupropion, respectively (Supplementary Table 6).

**Table 4.**
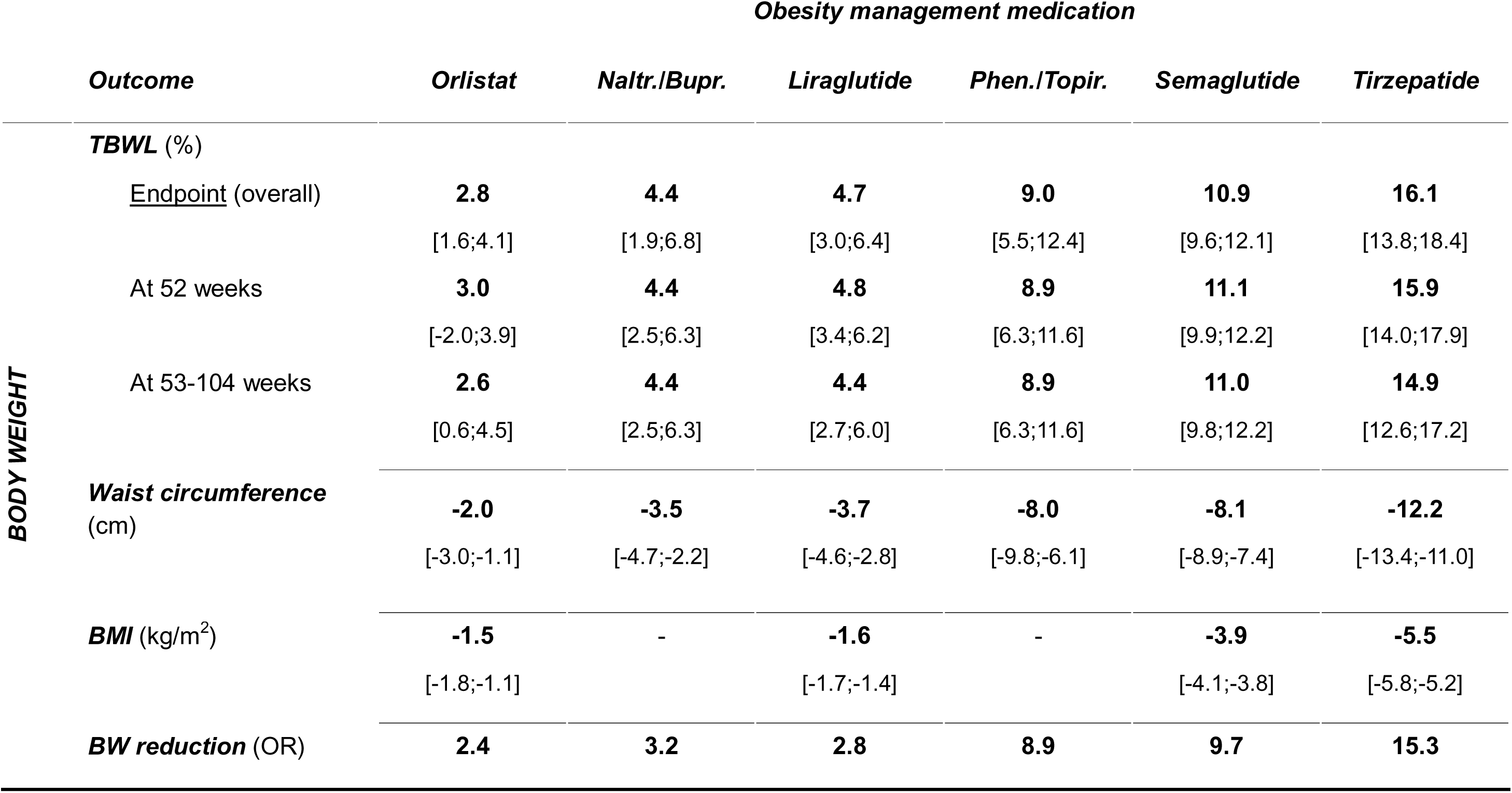
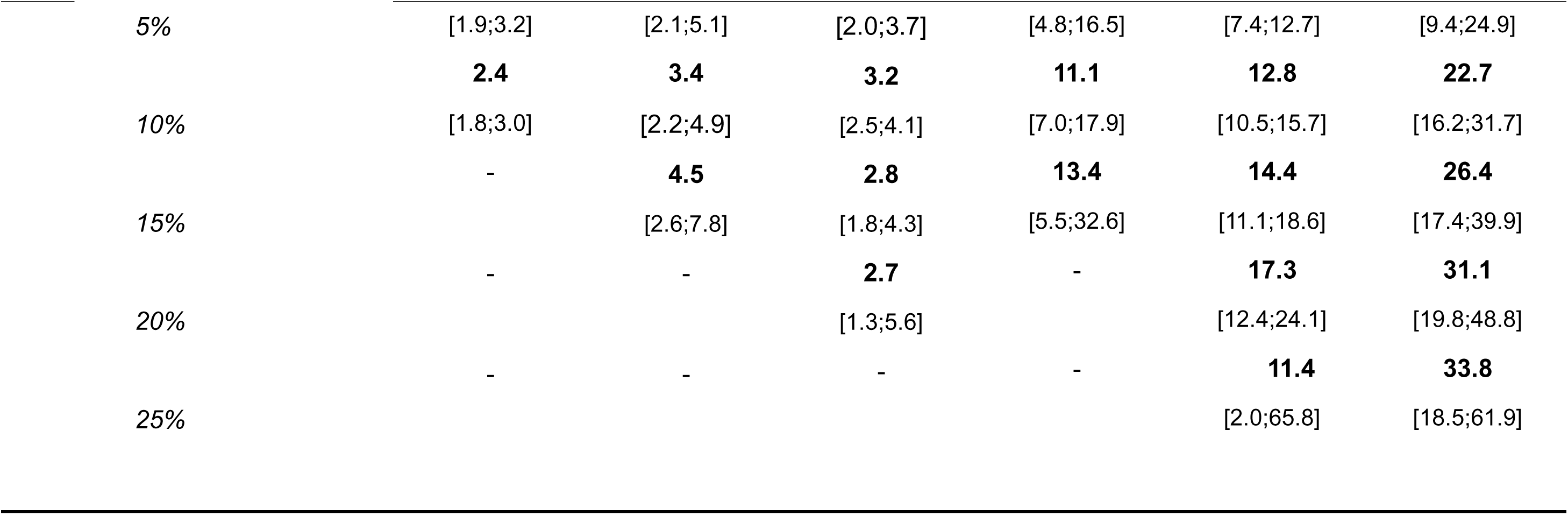
Effects of different OMM on body weight parameters (placebo-subtracted effect) in RCTs specifically designed for body weight reduction.

#### Heart failure

Three trials, two with semaglutide (Kosiborod 2023; Kosiborod 2024) and one with tirzepatide (Packer 2025), included patients with previously diagnosed HF; therefore, individual study results are presented. The effects of the two medications on TBWL% were not different from those observed in other non-cardiovascular outcome trials (Table 4). The risk of HHF was significantly reduced for both semaglutide and tirzepatide, together with increased disease-specific QoL questionnaire KCCQ scores and improved performance on the 6-minute walking test. No relevant effects on all-cause or cardiovascular mortality were observed for either medication (Table 4). The GRADE evaluation of the evidence retrieved for the primary endpoint (HHF, Supplementary Table 3) was rated as high for both tirzepatide and semaglutide (Supplementary Table 6).

#### Pre-diabetes

Two studies, one with liraglutide (le Roux 2017) and one with semaglutide (McGowan 2024), were specifically designed for patients with overweight/obesity and prediabetes, and two further studies, one with orlistat (Torgerson 2004) and one with tirzepatide (Jastreboff 2025), reported longer-term analyses in subgroups of patients with prediabetes; therefore, individual study results are presented. All four OMMs – orlistat, liraglutide, semaglutide, and tirzepatide – effectively reduced TBWL%, HbA1c, and FPG in patients with prediabetes.

Orlistat, liraglutide, and tirzepatide, but not semaglutide, were associated with a significant reduction in incident diabetes. Normoglycaemia restoration was more likely to be achieved in the intervention group for liraglutide, semaglutide, and tirzepatide, but not orlistat, compared with placebo (Table 4). The GRADE evaluation of the evidence retrieved for the primary endpoint (normoglycaemia restoration, Supplementary Table 3) was rated as high for all OMMs reporting this outcome, except for orlistat, for which the rating was low (Supplementary Table 6).

#### Type 2 diabetes

Fourteen trials were performed in patients with T2D (Table 4). Tirzepatide was associated with the greatest effect on TBWL%; semaglutide, tirzepatide, and liraglutide were all associated with significant effects on HbA1c and FPG. Naltrexone plus bupropion and orlistat did not have significant effects on FPG (Table 4). Fewer studies (n=4) examined effects on complete and partial diabetes remission rates; these study-specific estimates are presented, and all showed significant effects (Table 4). The GRADE evaluation of the evidence retrieved for the primary endpoint (diabetes remission, Supplementary Table 3) was rated as high for liraglutide, naltrexone plus bupropion, semaglutide, and tirzepatide (Supplementary Table 6).

#### Metabolic dysfunction-associated steatotic liver disease

Semaglutide (Loomba 2023; Sanyal 2025) and tirzepatide (Loomba 2024; Kadowaki 2025) were the only two OMMs assessed in patients with metabolic dysfunction-associated steatotic liver disease (MASLD). Tirzepatide resulted in a greater effect on TBWL% than semaglutide in these patients. Semaglutide reduced aspartate aminotransferase (AST), alanine aminotransferase (ALT), and liver stiffness, although the reductions in liver stiffness were not statistically significant. Tirzepatide also reduced AST, ALT, and liver stiffness, with the reduction in AST not reaching statistical significance. Both treatments were associated with significantly higher rates of metabolic dysfunction-associated steatohepatitis (MASH) remission, whereas only tirzepatide was associated with improvement of at least one fibrosis stage. Semaglutide and tirzepatide were both associated with a greater reduction in liver fat content compared with placebo (Table 4). The GRADE evaluation of the evidence retrieved for the primary endpoint (MASH remission, Supplementary Table 3) was rated as high and low for tirzepatide and semaglutide, respectively (Supplementary Table 6).

#### Obstructive sleep apnoea syndrome

One trial with tirzepatide enrolled patients living with obesity and OSAS (Malhotra 2024), reporting a reduction in the Apnoea-Hypopnoea Index (AHI), that is, the number of apnoeas and hypopnoeas per hour of sleep, and a greater percentage reduction in AHI (Table 4). The GRADE evaluation of the evidence retrieved for the primary endpoint (reduction of AHI episodes, Supplementary Table 3) was rated as moderate for tirzepatide (Supplementary Table 6).

#### Knee osteoarthritis

Semaglutide (Bliddal 2024) and liraglutide (Gudbergsen 2021) were assessed in patients with obesity and KOA. Semaglutide, but not liraglutide (Gudbergsen 2021), was capable of reducing knee pain and improving physical function assessed using the WOMAC scale, as shown in the individual studies (Table 4). The GRADE evaluation of the evidence retrieved for the primary endpoint (reduction of knee pain, Supplementary Table 3) was rated as moderate for semaglutide and low for liraglutide (Supplementary Table 6).

## Discussion

In this updated systematic review and meta-analysis, all included OMMs were associated with greater body weight reduction than placebo, but the magnitude of effect was not uniform. Tirzepatide and semaglutide produced the largest reductions in total body weight, and the direction of the three active-comparator trials was concordant with the network estimates, with liraglutide outperforming orlistat, semaglutide outperforming liraglutide, and tirzepatide outperforming semaglutide. Importantly, the clinical meaning of the evidence base has also broadened. Beyond weight loss, semaglutide and tirzepatide showed the strongest glycaemic effects, semaglutide was associated with lower risks of MACE and all-cause mortality, and dedicated complication-specific trials expanded the evidence base in heart failure with preserved ejection fraction, OSAS, KOA and MASLD. No OMMs were associated with an increased risk of serious adverse events.

These findings strengthen, rather than fundamentally change, the main conclusion of the 2025 EASO-associated review: pharmacological treatment of obesity is effective, but treatment choice should be individualized (McGowan 2025a; McGowan 2025b). What has changed is the maturity and scope of the evidence base. The earlier synthesis was driven predominantly by anthropometric and metabolic endpoints, with relatively limited direct active-comparator evidence and fewer dedicated trials focused on obesity-related complications. The present update preserves the hierarchy for weight-loss efficacy while adding direct comparative evidence from SURMOUNT-5 and substantially more complication-specific randomized data (Aronne 2025). As a result, the field is moving beyond the narrower question of which agent produces the greatest mean weight loss toward the more clinically relevant question of which agent is best matched to a given adiposity phenotype, dominant complication burden or treatment objective.

A clinically useful way to interpret these results is to distinguish weight-loss efficacy from evidence for complication modification. Tirzepatide currently appears to deliver the greatest average weight reduction and the strongest glycaemic effects. Semaglutide, however, has the most mature cardiovascular outcomes evidence in adults with overweight or obesity and established cardiovascular disease without diabetes because SELECT demonstrated a reduction in MACE. In dedicated HFpEF trials, semaglutide improved symptoms, physical function and body weight, whereas tirzepatide extended this evidence base by reducing the composite of cardiovascular death or worsening heart failure in SUMMIT. Tirzepatide also has dedicated randomized evidence in moderate-to-severe OSAS. In practice, this means the choice of OMMs may differ according to whether the dominant clinical priority is maximal total body weight loss, cardiovascular risk reduction, relief of heart-failure-related symptoms, reduction of sleep-disordered breathing, or improvement in metabolic liver disease. These findings should be interpreted with caution across complication domains, as several complication-specific outcomes are informed by only one or a small number of dedicated trials.

The liver findings are among the most important additions in this update. Compared with the previous review, the evidence base for histological liver outcomes is now substantially more informative. In the present synthesis, both semaglutide and tirzepatide were associated with higher rates of MASH resolution and reductions in liver fat content. For liver fibrosis, the evidence remains more limited and should be interpreted cautiously; however, the current dataset and the broader interpretation support semaglutide as having the more mature evidence base for fibrosis improvement at this stage, whereas evidence for tirzepatide is still less mature. These findings increase the clinical relevance of liver endpoints in obesity pharmacotherapy, while also underscoring the limits of direct cross-trial comparison across differences in disease stage, endpoint definition and trial design

Weight regain after treatment discontinuation deserves particular emphasis. Across withdrawal and extension studies, a substantial proportion of lost weight was regained after stopping liraglutide, semaglutide or tirzepatide. This is consistent with the understanding of obesity as a chronic, relapsing disease and argues against viewing OMMs as short, self-limited interventions for most patients. It also has practical implications for service design: treatment initiation should be accompanied by realistic discussion of likely duration of therapy, side-effect management, monitoring, and what happens if treatment must be interrupted because of cost, supply, intolerance or pregnancy planning. These issues are increasingly relevant to health-system implementation, as contemporary guidance is integrating OMMs into chronic obesity care pathways while still constraining access according to BMI, comorbidity and early treatment response.

The safety findings should also be interpreted in context. In this review, no OMMs were associated with a higher risk of serious adverse events, and no consistent signal emerged for suicide attempt, depression or anxiety. This is reassuring at the level of serious outcomes, but it should not be taken to mean that the evaluated OMMs are interchangeable from a tolerability perspective. Route of administration, gastrointestinal adverse effects, dose-escalation burden, contraindications, monitoring requirements and patient preferences remain clinically important determinants of persistence and real-world effectiveness, cost-effectiveness, and health system affordability. This is particularly relevant in obesity care, where long-term adherence is central to durability of benefit and where modest differences in efficacy may be outweighed by feasibility, acceptability or access in routine practice.

Like the previous review (McGowan 2025a), the present study has several limitations. First, the number of direct active-comparator trials remains small, and many complication-specific analyses are informed by single trials or by a small number of dedicated studies rather than by dense evidence networks. Second, trial populations, background lifestyle intervention, follow-up duration and definitions of remission or improvement were heterogeneous, particularly for quality-of-life, mental health, liver and body-composition outcomes. Third, long-term data beyond two to three years remain limited, and discontinuation studies remain comparatively sparse. Fourth, despite regulatory indications that extend to overweight with complications, no included trial had a mean baseline BMI below 30 kg/m², and only limited data were available in class III obesity. Finally, as with all NMAs, indirect comparisons rely on assumptions of transitivity and consistency that cannot be exhaustively tested for every clinical endpoint. These limitations argue for caution when making comparative inferences, especially for complication-specific outcomes supported by trial-specific designs.

The strengths of the present review are its comprehensive update of randomized evidence through the end of 2025, its focus on medications relevant to contemporary European practice, and its integration of anthropometric, metabolic, complication-specific, safety and GRADE-based assessments within a single framework. This is important because pharmacological obesity treatment is no longer defined only by mean percentage weight loss. The more clinically useful question is increasingly which medication best addresses the dominant clinical problem of a given patient while remaining feasible within the relevant health system. That interpretation is consistent with the current EASO framework and with recent National Institute for Health and Care Excellence (NICE) United Kingdom guidance, both of which move toward more individualized, complication-aware obesity care (NICE 2026). Future research should focus on long-term maintenance strategies, comparative effectiveness across phenotypes, sequencing or switching between agents, and standardized reporting of body composition, mental health and obesity-complication endpoints. More evidence is also needed in people with overweight and high complication burden, in severe obesity, and in populations underrepresented in current trials.

In conclusion, this updated network meta-analysis strengthens the original interpretation that OMMs should be selected according to the presence and profile of obesity-related complications, rather than on weight loss alone. Tirzepatide and semaglutide produced the greatest reductions in body weight, semaglutide remained the only agent associated with reduced MACE and all-cause mortality, and the updated evidence substantially expanded the liver domain. Taken together, these findings support a more individualized approach to obesity management pharmacotherapy based on expected weight-loss efficacy, complication profile, treatment goals, safety, patient preference and access.

## Data Availability

Data available upon request to the corresponding authors

## Acknowledgements

The authors thank Euan Woodward and the European Association for the Study of Obesity (EASO) for their support and research funding for this manuscript.

## Competing interests

A.C. has received speaking fees from Astra Zeneca, Boehringer-Ingelheim, Eli-Lilly, Novo Nordisk, Sanofi, Menarini; research grants from Eli Lilly, Novo Nordisk and Menarini; and is a member of the data monitoring committee of Boehringer Ingelheim. J.L.B. Advisory board member for Novo Nordisk (all fees paid to institution); representative of EASO on a Regeneron council (volunteer, no fees); speaker honoraria for participation in sponsored scientific sessions (all fees paid to institution); President-Elect of EASO. A.B. has received payment of honoraria from Astra Zeneca, Servier, Novo Nordisk, Eli Lilly and Novartis as speaker and/or member of advisory boards; Secretary of the Croatian Society of Obesity EASO ECN Board (Middle region), EASO COMs Working Group L.B. has payment of honoraria from Amgen, Boehringer Ingelheim, Bruno Farmaceutici, Eli-Lilly, Novo Nordisk, Pfizer, Recordati, Regeneron and Roche; speaker fees or educational activities from Pronokal and Rhythm Pharmaceuticals; Vice President Southern Region of EASO, member of the Standing Expert Panel for the endocrine-metabolic area of the National Guidelines System of the Italian Institute of Health (Istituto Superiore di Sanità). D.D. has received speaker and advisory board fees from Boehringer-Ingelheim, Eli-Lilly, Novo Nordisk, Astra Zeneca and research grants from Eli Lilly, Novo Nordisk and Boehringer Ingelheim. L.F. has received speaker and/or advisory fees from Novo Nordisk, Eli-Lilly, Zentiva, Stada, Vice-president Middle Region of EASO G.F. has received payment of honoraria from Eli Lilly, Novo Nordisk, Regeneron, Astra Zeneca and Marabou Foundation as speaker and/or member of advisory boards, and payment of honoraria as member of the OPEN Spain Initiative. G.G. has received research funding from Novo Nordisk, payment of honoraria as speaker (company educational seminars) from Novo Nordisk and Eli Lilly, and as Advisory Board member for Novo Nordisk (all fees paid to institution); co-chair of Basic and Discovery Science Working Group of EASO. J.G. is a shareholder of Elevate Access Ltd, which has received research and educational funding, including from the EASO. M.M. received speaking fees from Novo Nordisk and Eli Lilly, and consultancy fees from Novo Nordisk, Eli Lilly, and Boehringer Ingelheim. P.S. received payment of honoraria and consulting fees from Boehringer Ingelheim, Chiesi, Novo Nordisk, Eli Lilly, Pfizer, and Roche as a member of advisory boards. B.M.-T. has received grants from the EASO New Clinical Investigator Award 2024 and the EFSD Rising Star 2024, both supported by the Novo Nordisk Foundation. Matteo Monami has received speaking fees from Astra Zeneca, Bristol Myers Squibb, Boehringer-Ingelheim, Eli-Lilly, Merck, Novo Nordisk, Sanofi, and Novartis and research grants from Bristol Myers Squibb. V.Y. was engaged in advisory boards and lectures with Novo Nordisk, Eli Lilly, Rhythm and Regeneron; President of EASO. B.M. has received speaker and/or advisory fees from Novo Nordisk, Eli-Lilly, Astra Zeneca, Janssen, Pfizer, MSD and a research grant from Novo Nordisk; and is a shareholder of Reset Health.

## Notes

### Funding Statement

This study did not receive any funding

### Author Declarations

This is data from randomized clinical trial already published

## Reference

1. American Diabetes Association Professional Practice Committee for Obesity 2026. Pharmacologic treatment of obesity in adults: Standards of care in overweight and obesity. BMJ Open Diabetes Res Care, 13(Suppl 1), e005729. doi: 10.1136/bmjdrc-2025-005729. PMID: 41529914; PMCID: PMC12815056.

2. Aronne, L. J., Horn, D. B., le Roux, C. W., Ho, W., Falcon, B. L., Gomez Valderas, E., Das, S., Lee, C. J., Glass, L. C., Senyucel, C. & Dunn, J. P. 2025. Tirzepatide as Compared with Semaglutide for the Treatment of Obesity. N Engl J Med, 393, 26–36.

3. Aronne, L. J., Sattar, N., Horn, D. B., Bays, H. E., Wharton, S., Lin, W. Y., Ahmad, N. N., Zhang, S., Liao, R., Bunck, M. C., Jouravskaya, I. & Murphy, M. A. 2024. Continued Treatment With Tirzepatide for Maintenance of Weight Reduction in Adults With Obesity: The SURMOUNT-4 Randomized Clinical Trial. Jama, 331, 38–48.

4. Astrup, A., Carraro, R., Finer, N., Harper, A., Kunesova, M., Lean, M. E., Niskanen, L., Rasmussen, M. F., Rissanen, A., Rössner, S., Savolainen, M. J. & Van Gaal, L. 2012. Safety, tolerability and sustained weight loss over 2 years with the once-daily human GLP-1 analog, liraglutide. Int J Obes (Lond), 36, 843–54.

5. Bliddal, H., Bays, H., Czernichow, S., Uddén Hemmingsson, J., Hjelmesæth, J., Hoffmann Morville, T., Koroleva, A., Skov Neergaard, J., Vélez Sánchez, P., Wharton, S., Wizert, A. & Kristensen, L. E. 2024. Once-Weekly Semaglutide in Persons with Obesity and Knee Osteoarthritis. N Engl J Med, 391, 1573–1583.

6. Busetto, L., Dicker, D., Frühbeck, G., Halford, J. C. G., Sbraccia, P., Yumuk, V. & Goossens, G. H. 2024. A new framework for the diagnosis, staging and management of obesity in adults. Nat Med, 30, 2395–2399.

7. Ciudin, A. et al. 2026. Framework for the pharmacological treatment of obesity and its complications from the European Association for the Study of Obesity (EASO): 2026 update. Nat Med. NMED-C151239. DOI: 10.1038/s41591-026-04397-4.

8. G, S. meta: General Package for Meta-Analysis [Online]. [Accessed 27-03-2026 2026].

9. Garvey, W. T., Bluher, M., Osorto Contreras, C. K., Davies, M. J., Winning Lehmann, E., Pietilainen, K. H., Rubino, D., Sbraccia, P., Wadden, T., Zeuthen, N., Wilding, J. P. H. & Group, R. S. 2025. Coadministered Cagrilintide and Semaglutide in Adults with Overweight or Obesity. N Engl J Med, 393, 635–647.

10. Gudbergsen, H., Overgaard, A., Henriksen, M., Wæhrens, E. E., Bliddal, H., Christensen, R., Nielsen, S. M., Boesen, M., Knop, F. K., Astrup, A., Rasmussen, M. U., Bartholdy, C., Daugaard, C. L., Ellegaard, K., Heitmann, B. L., Bartels, E. M., Danneskiold-Samsøe, B. & Kristensen, L. E. 2021. Liraglutide after diet-induced weight loss for pain and weight control in knee osteoarthritis: a randomized controlled trial. Am J Clin Nutr, 113, 314–323.

11. Guyatt, G. H., Thorlund, K., Oxman, A. D., Walter, S. D., Patrick, D., Furukawa, T. A., Johnston, B. C., Karanicolas, P., Akl, E. A., Vist, G., Kunz, R., Brozek, J., Kupper, L. L., Martin, S. L., Meerpohl, J. J., Alonso-Coello, P., Christensen, R. & Schunemann, H. J. 2013. GRADE guidelines: 13. Preparing summary of findings tables and evidence profiles-continuous outcomes. J Clin Epidemiol, 66, 173–83.

12. Higgins, J. P., Altman, D. G., Gotzsche, P. C., Juni, P., Moher, D., Oxman, A. D., Savovic, J., Schulz, K. F., Weeks, L., Sterne, J. A., Cochrane Bias Methods, G. & Cochrane Statistical Methods, G. 2011. The Cochrane Collaboration’s tool for assessing risk of bias in randomised trials. Bmj, 343, d5928.

13. Jastreboff, A. M., Le Roux, C. W., Stefanski, A., Aronne, L. J., Halpern, B., Wharton, S., Wilding, J. P. H., Perreault, L., Zhang, S., Battula, R., Bunck, M. C., Ahmad, N. N. & Jouravskaya, I. 2024. Tirzepatide for Obesity Treatment and Diabetes Prevention. N Engl J Med.

14. Jastreboff, A. M., Le Roux, C. W., Stefanski, A., Aronne, L. J., Halpern, B., Wharton, S., Wilding, J. P. H., Perreault, L., Zhang, S., Battula, R., Bunck, M. C., Ahmad, N. N. & Jouravskaya, I. 2025. Tirzepatide for Obesity Treatment and Diabetes Prevention. N Engl J Med, 392, 958–971.

15. Kadowaki, T., Isendahl, J., Khalid, U., Lee, S. Y., Nishida, T., Ogawa, W., Tobe, K., Yamauchi, T. & Lim, S. 2022. Semaglutide once a week in adults with overweight or obesity, with or without type 2 diabetes in an east Asian population (STEP 6): a randomised, double-blind, double-dummy, placebo-controlled, phase 3a trial. Lancet Diabetes Endocrinol, 10, 193–206.

16. Kadowaki, T., Nishida, T., Ogawa, W., Overvad, M., Tobe, K. & Yamauchi, T. 2025. Effect of once-weekly subcutaneous semaglutide on abdominal visceral fat area in Japanese adults with overweight and obesity: A post hoc analysis of the STEP 6 trial. Obes Res Clin Pract, 19, 146–153.

17. Karhunen, L., Franssila-Kallunki, A., Rissanen, P., Valve, R., Kolehmainen, M., Rissanen, A. & Uusitupa, M. 2000. Effect of orlistat treatment on body composition and resting energy expenditure during a two-year weight-reduction programme in obese Finns. Int J Obes Relat Metab Disord, 24, 1567–72.

18. Kosiborod, M. N., Abildstrøm, S. Z., Borlaug, B. A., Butler, J., Rasmussen, S., Davies, M., Hovingh, G. K., Kitzman, D. W., Lindegaard, M. L., Møller, D. V., Shah, S. J., Treppendahl, M. B., Verma, S., Abhayaratna, W., Ahmed, F. Z., Chopra, V., Ezekowitz, J., Fu, M., Ito, H., Lelonek, M., Melenovsky, V., Merkely, B., Núñez, J., Perna, E., Schou, M., Senni, M., Sharma, K., van der Meer, P., von Lewinski, D., Wolf, D. & Petrie, M. C. 2023. Semaglutide in Patients with Heart Failure with Preserved Ejection Fraction and Obesity. N Engl J Med, 389, 1069–1084.

19. Kosiborod, M. N., Petrie, M. C., Borlaug, B. A., Butler, J., Davies, M. J., Hovingh, G. K., Kitzman, D. W., Møller, D. V., Treppendahl, M. B., Verma, S., Jensen, T. J., Liisberg, K., Lindegaard, M. L., Abhayaratna, W., Ahmed, F. Z., Ben-Gal, T., Chopra, V., Ezekowitz, J. A., Fu, M., Ito, H., Lelonek, M., Melenovský, V., Merkely, B., Núñez, J., Perna, E., Schou, M., Senni, M., Sharma, K., van der Meer, P., von Lewinski, D., Wolf, D. & Shah, S. J. 2024. Semaglutide in Patients with Obesity-Related Heart Failure and Type 2 Diabetes. N Engl J Med, 390, 1394–1407.

20. Le Roux, C. W., Astrup, A., Fujioka, K., Greenway, F., Lau, D. C. W., van Gaal, L., Ortiz, R. V., Wilding, J. P. H., Skjøth, T. V., Manning, L. S. & PI-Sunyer, X. 2017. 3 years of liraglutide versus placebo for type 2 diabetes risk reduction and weight management in individuals with prediabetes: a randomised, double-blind trial. Lancet, 389, 1399–1409.

21. Lincoff, A. M., Brown-Frandsen, K., Colhoun, H. M., Deanfield, J., Emerson, S. S., Esbjerg, S., Hardt-Lindberg, S., Hovingh, G. K., Kahn, S. E., Kushner, R. F., Lingvay, I., Oral, T. K., Michelsen, M. M., Plutzky, J., Tornøe, C. W. & Ryan, D. H. 2023. Semaglutide and Cardiovascular Outcomes in Obesity without Diabetes. N Engl J Med, 389, 2221–2232.

22. Loomba, R., Abdelmalek, M. F., Armstrong, M. J., Jara, M., Kjær, M. S., Krarup, N., Lawitz, E., Ratziu, V., Sanyal, A. J., Schattenberg, J. M. & Newsome, P. N. 2023. Semaglutide 2·4 mg once weekly in patients with non-alcoholic steatohepatitis-related cirrhosis: a randomised, placebo-controlled phase 2 trial. Lancet Gastroenterol Hepatol, 8, 511–522.

23. Loomba, R., Hartman, M. L., Lawitz, E. J., Vuppalanchi, R., Boursier, J., Bugianesi, E., Yoneda, M., Behling, C., Cummings, O. W., Tang, Y., Brouwers, B., Robins, D. A., Nikooie, A., Bunck, M. C., Haupt, A., Sanyal, A. J. & Investigators, S.-N. 2024. Tirzepatide for Metabolic Dysfunction-Associated Steatohepatitis with Liver Fibrosis. N Engl J Med, 391, 299–310.

24. Lundgren, J. R., Janus, C., Jensen, S. B. K., Juhl, C. R., Olsen, L. M., Christensen, R. M., Svane, M. S., Bandholm, T., Bojsen-Møller, K. N., Blond, M. B., Jensen, J. B., Stallknecht, B. M., Holst, J. J., Madsbad, S. & Torekov, S. S. 2021. Healthy Weight Loss Maintenance with Exercise, Liraglutide, or Both Combined. N Engl J Med, 384, 1719–1730.

25. Malhotra, A., Grunstein, R. R., Fietze, I., Weaver, T. E., Redline, S., Azarbarzin, A., Sands, S. A., Schwab, R. J., Dunn, J. P., Chakladar, S., Bunck, M. C. & Bednarik, J. 2024. Tirzepatide for the Treatment of Obstructive Sleep Apnea and Obesity. N Engl J Med.

26. Mcgowan, B., Ciudin, A., Baker, J. L. et al. 2025a. A systematic review and meta-analysis of the efficacy and safety of pharmacological treatments for obesity in adults. Nat Med, 31, 3317–3329. 10.1038/s41591-025-03978-z.

27. Mcgowan, B., Ciudin, A., Baker, J. L. et al. 2025b. Framework for the pharmacological treatment of obesity and its complications from the European Association for the Study of Obesity (EASO). Nat Med, 31, 3229–3232. 10.1038/s41591-025-03765-w.

28. Mcgowan, B. M., Bruun, J. M., Capehorn, M., Pedersen, S. D., Pietiläinen, K. H., Muniraju, H. A. K., Quiroga, M., Varbo, A. & Lau, D. C. W. 2024. Efficacy and safety of once-weekly semaglutide 2·4 mg versus placebo in people with obesity and prediabetes (STEP 10): a randomised, double-blind, placebo-controlled, multicentre phase 3 trial. Lancet Diabetes Endocrinol, 12, 631–642.

29. Nagi, M. A., Ahmed, H., Rezq, M. A. A., Sangroongruangsri, S., Chaikledkaew, U., Almalki, Z. & Thavorncharoensap, M. 2024. Economic costs of obesity: a systematic review. Int J Obes (Lond), 48, 33–43. doi: 10.1038/s41366-023-01398-y. Epub 2023 Oct 26. PMID: 37884664.

30. Nissen, S. E., Wolski, K. E., Prcela, L., Wadden, T., Buse, J. B., Bakris, G., Perez, A. & Smith, S. R. 2016. Effect of Naltrexone-Bupropion on Major Adverse Cardiovascular Events in Overweight and Obese Patients With Cardiovascular Risk Factors: A Randomized Clinical Trial. Jama, 315, 990–1004.

31. Packer, M., Zile, M. R., Kramer, C. M., Baum, S. J., Litwin, S. E., Menon, V., Ge, J., Weerakkody, G. J., Ou, Y., Bunck, M. C., Hurt, K. C., Murakami, M. & Borlaug, B. A. 2025. Tirzepatide for Heart Failure with Preserved Ejection Fraction and Obesity. N Engl J Med, 392, 427–437.

32. Richelsen, B., Tonstad, S., Rössner, S., Toubro, S., Niskanen, L., Madsbad, S., Mustajoki, P. & Rissanen, A. 2007. Effect of orlistat on weight regain and cardiovascular risk factors following a very-low-energy diet in abdominally obese patients: a 3-year randomized, placebo-controlled study. Diabetes Care, 30, 27–32.

33. Rubino, D. M., Greenway, F. L., Khalid, U., O’neil, P. M., Rosenstock, J., Sørrig, R., Wadden, T. A., Wizert, A. & Garvey, W. T. 2022. Effect of Weekly Subcutaneous Semaglutide vs Daily Liraglutide on Body Weight in Adults With Overweight or Obesity Without Diabetes: The STEP 8 Randomized Clinical Trial. Jama, 327, 138–150.

34. Rucker G, K. U. netmeta: Network Meta-Analysis using Frequentist Methods [Online]. [Accessed 27-03-2026 2026].

35. Sanyal, A. J., Newsome, P. N., Kliers, I., Østergaard, L. H., Long, M. T., Kjær, M. S., Cali, A. M. G., Bugianesi, E., Rinella, M. E., Roden, M. & Ratziu, V. 2025. Phase 3 Trial of Semaglutide in Metabolic Dysfunction-Associated Steatohepatitis. N Engl J Med, 392, 2089–2099.

36. Swinburn, B. A., Carey, D., Hills, A. P., Hooper, M., Marks, S., Proietto, J., Strauss, B. J., Sullivan, D., Welborn, T. A. & Caterson, I. D. 2005. Effect of orlistat on cardiovascular disease risk in obese adults. Diabetes Obes Metab, 7, 254–62.

37. Torgerson, J. S., Hauptman, J., Boldrin, M. N. & Sjöström, L. 2004. XENical in the prevention of diabetes in obese subjects (XENDOS) study: a randomized study of orlistat as an adjunct to lifestyle changes for the prevention of type 2 diabetes in obese patients. Diabetes Care, 27, 155–61.

38. Valera-Mora, M. E., Simeoni, B., Gagliardi, L., Scarfone, A., Nanni, G., Castagneto, M., Manco, M., Mingrone, G. & Ferrannini, E. 2005. Predictors of weight loss and reversal of comorbidities in malabsorptive bariatric surgery. Am J Clin Nutr, 81, 1292–7.

39. Wharton, S., Freitas, P., Hjelmesaeth, J., Kabisch, M., Kandler, K., Lingvay, I., Quiroga, M., Rosenstock, J., Garvey, W. T. & Group, S. U. T. 2025. Once-weekly semaglutide 7.2 mg in adults with obesity (STEP UP): a randomised, controlled, phase 3b trial. Lancet Diabetes Endocrinol, 13, 949–963.

40. WHO 2025. Fact Sheets – Obesity and overweight. Available at: https://www.who.int/en/news-room/fact-sheets/detail/obesity-and-overweight. Accessed: 15 March 2026.

41. Wilding, J. P. H., Batterham, R. L., Calanna, S., Davies, M., van Gaal, L. F., Lingvay, I., Mcgowan, B. M., Rosenstock, J., Tran, M. T. D., Wadden, T. A., Wharton, S., Yokote, K., Zeuthen, N. & Kushner, R. F. 2021. Once-Weekly Semaglutide in Adults with Overweight or Obesity. N Engl J Med, 384, 989–1002.

42. Wilding, J. P. H., Batterham, R. L., Davies, M., van Gaal, L. F., Kandler, K., Konakli, K., Lingvay, I., Mcgowan, B. M., Oral, T. K., Rosenstock, J., Wadden, T. A., Wharton, S., Yokote, K. & Kushner, R. F. 2022. Weight regain and cardiometabolic effects after withdrawal of semaglutide: The STEP 1 trial extension. Diabetes Obes Metab, 24, 1553–1564.

43. World Obesity Federation 2025. World Obesity Atlas 2025. Available at: https://www.worldobesity.org/resources/resource-library/world-obesity-atlas-2025. Accessed: 15 March 2026.

